# Integrative screening identifies functional variants and VNTRs underlying GWAS signals at the 5p15.33 multi-cancer susceptibility locus

**DOI:** 10.64898/2026.03.03.26347427

**Authors:** Aidan O’Brien, Hyunkyung Kong, Harsh Patel, Michelle Ho, Minal B. Patel, Jun Zhong, Mai Xu, Brenen W. Papenberg, Katelyn E. Connelly, Irene Collins, Rebecca Hennessey, Rohit Thakur, Hayley Sowards, Karen Funderburk, Thong Luong, Oscar Florez-Vargas, Timothy Myers, Ashley Jermusyk, Bryan Gorman, Wen Luo, Kristine Jones, Sudipto Das, The Pancreatic Cancer Cohort and Case-Control Consortia (PanScan/PanC4), Melanoma Meta-Analysis Consortium, International Lung Cancer Consortium (ILCCO), Qing Lan, Nathaniel Rothman, James D. McKay, Rayjean J. Hung, Christopher I. Amos, Mark M. Iles, Stella Koutros, Maria Teresa Landi, Matthew H. Law, Rachael Z. Stolzenberg-Solomon, Brian M Wolpin, Manal Hassan, Alison P. Klein, Samuel O Antwi, Nick Orr, Stephen J. Chanock, Sara Lindström, Jason W. Hoskins, Marc-Henri Stern, Thorkell Andresson, Jianxin Shi, Ludmila Prokunina-Olsson, Jiyeon Choi, Kevin M. Brown, Laufey T. Amundadottir

## Abstract

Chromosome 5p15.33 harbors several independent association signals which demonstrate antagonistic pleiotropy across cancer types, with causal mechanisms largely unresolved. To identify functional variants and enhancer elements at this locus, we performed statistical fine-mapping followed by massively parallel reporter assays (MPRA) and proliferation based CRISPRi screens. This approach identified eight multi-cancer functional variants (MCFVs) across three GWAS signals. Targeting rs421629 (part of the *CLPTM1L* signal marked by rs465498) with CRISPRi revealed opposing effects on *TERT* expression in pancreatic versus lung cancer cells, consistent with the antagonistic pleiotropy observed for this signal. Furthermore, CRISPRi nominated an intronic *CLPTM1L* variable number tandem repeat (VNTR) as a potent enhancer. Long-read sequencing established VNTR polymorphisms as potential causal variants for the rs465498 signal. We showed that Hippo-pathway transcription factors mediate VNTR enhancer activity in lung and pancreatic cancer cells. Together, these findings indicate that cancer susceptibility at 5p15.33 may be mediated by both SNPs and VNTRs and provide an integrated framework for resolving complex pleiotropic loci.

## Introduction

While genome-wide association studies (GWAS) have identified hundreds of common risk loci for human cancers, functional characterization of the association signals has lagged behind their identification^1^. Many such loci are thought to be underpinned by causal variants which influence allele-specific *cis*-regulation of nearby target genes^2,3^. Chromosome 5p15.33 is one of the most replicated cancer susceptibility loci, where at least 10 distinct signals have been identified across as many cancer types^4,5^. Successive multi-cancer fine-mapping analyses by Wang and colleagues in 2014^4^ and Chen and colleagues in 2021^5^ have applied the Association Analysis for SubSETs (ASSET)^6^ method to GWAS datasets at the 5p15.33 locus, noting considerable genetic correlation between cancer types. Intriguingly, signals at 5p15.33 demonstrate antagonistic pleiotropy, with risk alleles for one cancer being protective for other cancer types^4,5,7^.

At 5p15.33, both *TERT* and *CLPTM1L* are plausible target genes with established roles in tumor development and progression. *TERT* encodes the catalytic subunit of telomerase reverse transcriptase, which maintains telomere length and protects chromosomes from replication-associated erosion. Telomerase activation via upregulation of *TERT* is observed in over 90% of cancers^8–10^. Furthermore, *TERT* represents an established cancer susceptibility gene via both rare and common germline promoter polymorphisms^11–16^. Somatic *TERT* promoter mutations are prognostic indicators for multiple cancers^17–21^, and act synergistically with proximal germline polymorphisms (rs2853669) in melanoma^22^, bladder cancer^18,23^, and glioma^21,24^. Notably, cancers such as pancreatic ductal adenocarcinoma (PDAC) exhibit activated telomerase activity^25^ despite lacking canonical *TERT* promoter mutations^26^, suggesting alternate *cis*-regulatory activating mechanisms. *CLPTM1L* encodes a transmembrane lipid scramblase which is overexpressed in pancreatic, ovarian, and lung cancers and contributes to RAS-dependent transformation^27^, acting as an anti-apoptotic factor upon endoplasmic reticulum stress^28–31^. Recent functional analyses^32,33^ and transcriptome-wide association studies^34^ further implicate *CLPTM1L* expression as a potential mediator of risk for uveal melanoma and pancreatic cancer, respectively. Together, these observations raise the possibility that 5p15.33 risk alleles converge through expression of *TERT*, *CLPTM1L*, or both, motivating a high-throughput, cross-cancer functional screen of cancers associated with the 5p15.33 locus using methodologies sensitive to downstream effects on each gene.

Post-GWAS studies focusing on specific association signals and cancer types have characterized *cis*-regulatory effects of functional sequence variants on both *TERT* and *CLPTM1L* as effector genes^7,12,32,35–40^. More recently, data presented by Florez-Vargas and colleagues^41^ suggests that variable number tandem repeats (VNTRs) may represent an important class of credible causal variants (CCVs) at 5p15.33^42–44^, indicating that SNP-centric models of GWAS causality may overlook more cryptic classes of functional variants. With no extensive cross-cancer screen of regulatory elements and CCVs performed for the larger region to date, we set out to comprehensively characterize *cis*-regulatory elements and CCVs to better understand the mechanisms underpinning cancer risk and pleiotropy at this important multi-cancer susceptibility locus.

To systematically characterize functional variants and *cis*-regulatory elements important for putative 5p15.33 effector genes, we performed an integrative functional screen at this locus. First, we fine-mapped GWAS datasets from melanoma, lung, pancreatic, and bladder cancers and compiled correlated variants from previously published signals to derive a panel of cross-cancer CCVs. We then implemented a massively parallel reporter assay (MPRA) to assess allele-specific regulatory activity for CCVs in cell lines representative of the four fine-mapped cancers. Next, we agnostically identified *cis*-regulatory elements using a tiled CRISPR-inhibition (CRISPRi) proliferation screen in cell lines from the same four cancer types. By integrating MPRA and CRISPRi results, we derived multi-cancer functional variants (MCFV) which mediate their effect through *cis*-regulation of *TERT* and/or *CLPTM1L* across cancer types. In addition, CRISPRi screening nominated a variable number tandem repeat (VNTR) within intron 9 of *CLPTM1L* as a potent enhancer element. Long-read sequencing, followed by imputation of VNTR alleles into a pancreatic cancer GWAS dataset, revealed that polymorphisms of this element contribute to the credible set of variants underlying the rs465498 signal at *CLPTM1L*. Subsequent functional analyses revealed VNTR-mediated regulation of both *TERT* and *CLPTM1L* through Hippo signaling pathway transcription factors.

Collectively, our work applies and extends a generalizable framework for integrative high-throughput functional screens to assess causal regulatory mechanisms at complex GWAS loci. By expanding our scope to repeat polymorphisms, we demonstrate that VNTRs and SNPs may jointly underlie cancer susceptibility signals through *cis*-regulation of both *TERT* and *CLPTM1L*. In doing so, our work bridges a gap between statistical genetics and functional biology, providing new insights into the molecular basis of cancer heritability at one of the most recurrent and complex cancer susceptibility loci identified through GWAS.

## Results

### Fine-mapping reveals multiple credible sets at 5p15.33

To comprehensively compile credible causal variants (CCVs) for massively parallel reporter assay (MPRA) analysis, we performed ancestry-specific fine-mapping of GWAS summary statistics from four 5p15.33-implicated cancer types in populations of European ancestry using SuSiE^45^ to derive 95% credible causal sets (PDAC: 9,040 cases and 12,496 control subjects^46^, melanoma: 36,760 cases and 375,188 controls^47^, bladder cancer: 13,790 cases and 343,502 controls^48^ and lung adenocarcinoma: 29,266 cases and 56,450 controls^49^). We also assessed a lung adenocarcinoma GWAS of 21,658 cases and 150,676 controls of East Asian^50^ (EAS) ancestry (**Table 1**, **Fig. 1, Supplementary Figs. 1&2)**. These credible sets were denoted by their lead variant (highest posterior inclusion probability, PIP) and assigned to individual pleiotropic ASSET signals reported by Chen and colleagues^5^ based on pairwise LD between credible set lead variants and ASSET tag variants (*r*^2^ ≥ 0.6, 1000 Genomes Project Phase 3, 1kGP p3^51^, **Fig. 2, Methods**).

**Fig. 1:**
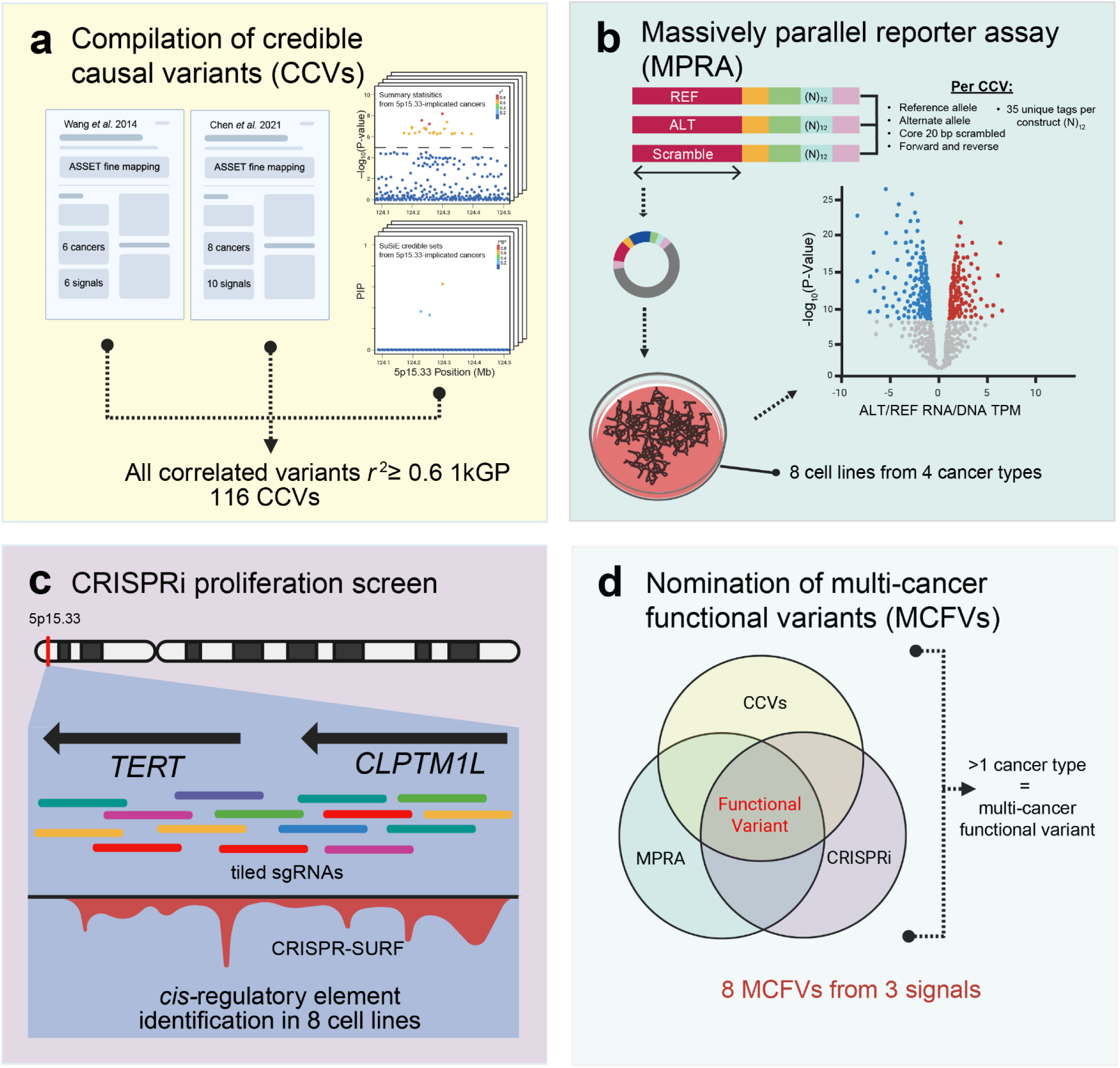
Integrative screening strategy for identification of 5p15.33 multi-cancer functional variants. **a**, To collate credible causal variants (CCVs) at 5p15.33 for MPRA, we initially compiled signal-tagging lead variants from two multi-cancer fine-mapping studies (Chen et al. 2021^5^; Wang et al. 2014^4^) along with credible set-tagging variants resulting from SuSiE fine-mapping of GWAS summary statistics from four 5p15.33-implicated cancers. We carried forward lead/tagging variants, additional SuSiE credible-set CCVs, and ancestry-matched LD-correlated variants (1kGP Phase 3; *r*^2^ ≥ 0.6 for downstream analyses, total 116 CCVs). **b,** Fine mapped CCVs were subject to MPRA to characterize allelic effects on transcriptional output across eight cell lines representative of the four fine mapped cancer types. **c,** Additionally, we assayed across the larger locus to nominate *cis*-regulatory elements using a tiled CRISPRi proliferation screen across eight cell lines spanning four cancer types. **d,** We then integrated results across cancer types to nominate eight multi-cancer functional variants (MCFVs) across three distinct signals.

**Fig. 2:**
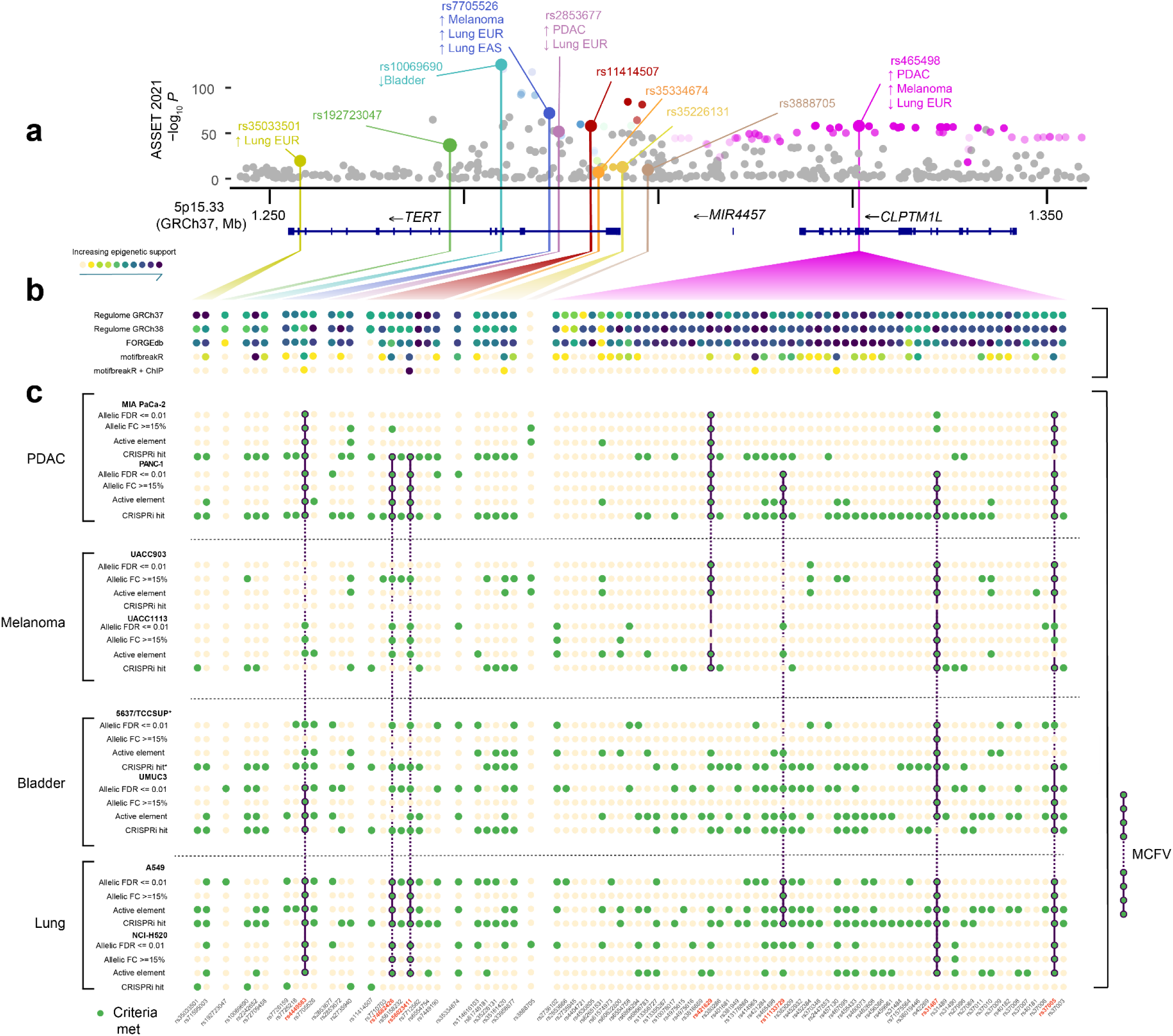
Functional screening reveals multi-cancer functional variants (MCFVs) across 5p15.33. **a,** CCVs were stratified by pairwise LD *r*^2^ ≥ 0.6 relative to signal-tagging variants reported by Chen *et al.* 2021^5^. ASSET signals were annotated with a cancer type if SuSiE fine-mapping yielded a credible set for a given cancer corresponding to that signal using the same LD threshold. Arrows assigned to each credible set indicate the relative direction of effect for each fine-mapped cancer (↑ risk increasing, ↓ protective) (top panel). **b,** CCVs were annotated with RegulomeDB and FORGEdb scores and motifbreakR predictions of transcription factor motif disruption. MotifbreakR results were further annotated for concordant TF-ChIP-seq data from the ReMap2022 database^93^ (motifbreakR + ChIP) (middle panel, full annotations in **Supplementary Table 3**). **c,** For each CCV, binary indicators were assigned per cell line based on MPRA allelic activity (FDR-significant effect, ≥ 15% absolute fold change, and active-element status) and overlap with proliferation-essential elements from CRISPRi. CCVs were nominated as screening hits when MPRA-functional status was supported by CRISPRi within cell lines from the same cancer type (green filled circles). CCVs were further categorized as multi-cancer functional variants (MCFVs) when screening hit status was observed in more than one cancer type (green filled connected circles with black outline; rsIDs highlighted in red, bottom panels). Functional support was strongest for CCVs mapping to the rs465498 signal (four MCFVs) over *CLPTM1L* and the rs11414507 signals (two MCFVs) over *TERT*. rs62332591 was also nominated as a MCFV but was not correlated at *r*^2^ ≥ 0.6 to an ASSET signal (and not shown in this figure).

**Table 1:** Credible sets identified via fine-mapping at 5p15.33 across four cancer types.

| GWAS Dataset | Most significant 5p15.33 SNP | | | | | | | SuSiE credible sets | | Chen et al. 2021 SNP ($r^2$ 1kGP EUR) |
| --- | --- | --- | --- | --- | --- | --- | --- | --- | --- | --- |
|  | rsID | chr5 Position (GRCh37) | chr5 Position (GRCh38) | Effect Allele | Non-Effect Allele | Odds Ratio | P-Value | CS-tagging CCV | nSNPs |  |
| PDAC<sup>46</sup> | rs31490 | 1344458 | 1344343 | G | A | 0.84 | 6.02x10<sup>-17</sup> | rs31490 | 19 | rs465498 (0.96) |
|  |  |  |  |  |  |  |  | rs2735940 | 2 | rs2853677 (0.62) |
| Melanoma<sup>47</sup> | rs13178866 | 1323212 | 1323097 | C | T | 0.88 | 1.10x10<sup>-43</sup> | rs13178866 | 11 | rs465498 (0.98) |
|  |  |  |  |  |  |  |  | rs2736100 | 1 | rs7705526 (0.51) |
|  |  |  |  |  |  |  |  | rs2853667 | 1 | - |
| Lung EUR<sup>49</sup> | rs7705526 | 1285974 | 1285859 | G | A | 1.25 | 3.80x10<sup>-35</sup> | rs7705526 | 1 | rs7705526 |
|  |  |  |  |  |  |  |  | rs6554758 | 2 | rs465498 (0.77) |
|  |  |  |  |  |  |  |  | rs2853677 | 1 | rs2853677 |
|  |  |  |  |  |  |  |  | rs35033501 | 2 | rs35033501 |
| Lung EAS<sup>50</sup> | rs2736100 | 1286516 | 1286401 | C | A | 0.75 | 7.92x10<sup>-58</sup> | rs2736100 | 1 | rs7705526 (0.52, 0.859 EAS) |
| Bladder<sup>48</sup> | rs2242652 | 1280028 | 1279913 | A | G | 1.08 | 4.06x10<sup>-15</sup> | rs10069690 | 1 | rs10069690 |

Across studies, SuSiE resolved 11 cancer-specific credible sets (ranging from 1–19 variants; **Table 1, Supplementary Fig. 2**), with 10/11 credible sets recapitulating previously reported ASSET signals (**Table 1**). Lung EUR yielded four credible sets, consistent with previously reported signals^49^. Lung EAS, by contrast, yielded a single credible set marked by rs2736100. Fine-mapping for bladder cancer resulted in a single credible set mapping to the signal marked by rs10069690. Additionally, SuSiE fine-mapping nominated a melanoma signal marked by rs2853667 that was not captured by ASSET (**Table 1**). The rs465498-tagged signal mapping across *CLPTM1L,* notably yielding a larger number of CCVs relative to other signals, was supported in PDAC, melanoma, and lung EUR GWAS, with antagonistic pleiotropy consistent with previous analyses (**Supplementary Fig. 2f**). SuSiE did not return a credible set for several previously reported association signals (PDAC rs2736098^52^, lung EAS rs1316780 and rs62332591^50^; **Supplementary Fig. 2**.). In two instances the best-matching ASSET signal was supported only by modest LD with these SNPs (PDAC rs2735940 with rs2853677, *r*² = 0.62; melanoma rs2736100 with rs7705526, *r*² = 0.51; **Table 1**).

To ensure comprehensive coverage for our MPRA design, we supplemented ASSET signals and cancer-specific CCVs with ancestry-matched proxy variants from 1kGP p3 (*r*² ≥ 0.6) to derive a final MPRA panel of 116 5p15.33 CCVs (**Fig. 1a; Supplementary Fig. 1, Supplementary Table 1**). Given considerable cross-correlation of SuSiE credible sets (**Supplementary Fig. 3**), subsequent functional annotation of CCVs was stratified by ASSET signal, with SuSiE results used supplementarily to ensure comprehensive functional annotation across pleiotropic signals.

### MPRA identifies functional *cis*-regulatory CCVs

To assess allele-specific *cis*-regulatory activity for each of the 116 fine-mapped CCVs, we utilized an MPRA strategy based on previously established workflows^53–56^ across eight cancer cell lines representative of the four fine-mapped cancers (pancreas: PANC-1 and MIA PaCa-2; melanoma: UACC903 and UACC1113; bladder: 5637 and UM-UC-3; lung: A549 and NCI-H520). After quality control (see **Methods, Supplementary Fig. 4**), 61.21% of tested CCVs demonstrated a significant allelic effect (False Discovery Rate, FDR ≤ 0.01) in at least one cell line (**Supplementary Table 2**), although effect sizes were generally modest (median allelic log_2_ fold change for significant CCVs was 0.067, **Supplementary Fig. 5**). We therefore prioritized CCVs using additional stringency metrics in a step-wise manner: (i) active element status for the 145 bp sequence harboring each variant determined by divergence of either allele from a distribution of scrambled controls (n = 79 of 109 CCVs passing MPRA design and QC; see **Methods; Supplementary Fig. 6**) (ii) FDR ≤ 0.01 for allelic effects (n = 71), (iii) absolute allelic Δ RNA/DNA TPM ≥ 15% (n = 13). Variants meeting all three criteria in at least one cell line were designated MPRA-hits (n = 13; **Table 2**). Effect sizes were concordant with independent MPRA experiments of overlapping CCVs in UACC903 cells by Choi and colleagues^55^ (Pearson’s *r* = 0.88, n = 5), and lung cancer cell lines by Long and colleagues^54^ (A549 *r* = 0.84, NCI-H520 *r* = 0.82, n = 17). MPRA-functional CCVs mapped to five ASSET signals (marked by rs465498: n = 7 CCVs; rs11414507: 2; rs7705526: 1; rs7726159: 2; rs451360: 1; **Supplementary Table 3).** We annotated CCVs with functional scores from RegulomeDB^57^, FORGEdb^58^, and assessed disruption/creation of transcription factor binding sites with motifbreakR^59^ (**Supplementary Table 4**), but did not observe robust enrichment of MPRA hits among higher-scoring or motif-disrupting candidates (see **Supplementary Results**), underscoring the value of direct functional screening at this locus.

**Table 2:**
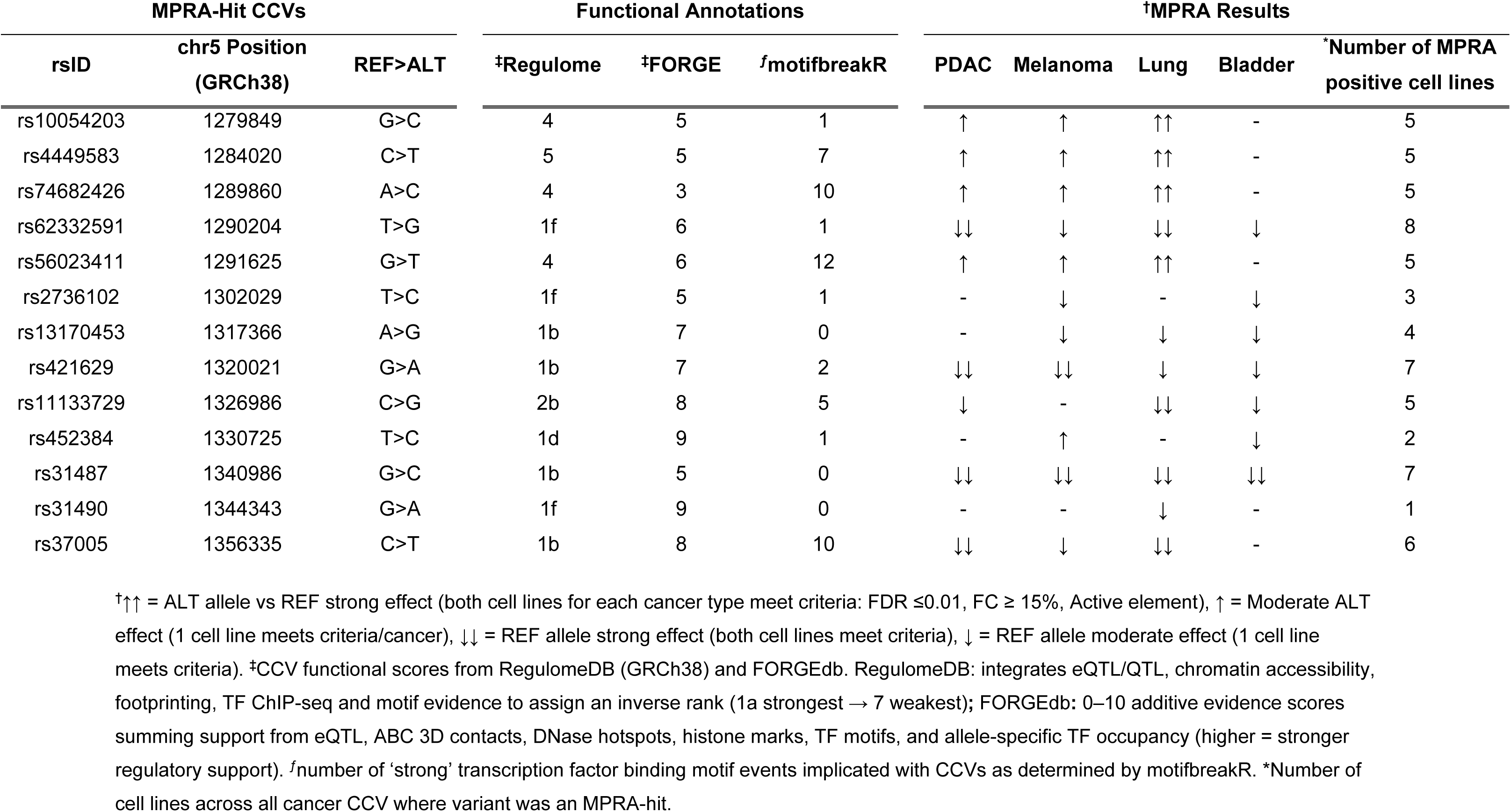
5p15.33 MPRA results and functional annotations.

### A tiled CRISPRi screen at 5p15.33 supported MPRA-hits and revealed additional regulatory elements

Because episomal MPRA constructs are not integrated into the endogenous chromatin context, we complemented this approach with an agnostic, tiled CRISPRi proliferation screen of 25,778 sgRNAs across this locus (chr5:1,233,287-1,380,000, GRCh37) ensuring robust coverage of CCVs across the sgRNA pool (**Supplementary Table 5, Supplementary Results**). We used representative cell lines from the same four cancer types engineered to stably express dCas9-KRAB-ZIM3, retaining seven of the eight lines described above and substituting TCCSUP for 5637 for bladder cancer (see **Methods**). After confirming both *TERT* and *CLPTM1L* as proliferation-essential genes in these cell lines (consistent with data from DepMap^60^, knockdown data not presented), CRISPRi screens were carried out over 25 cumulative cell doublings with sampling at T_0_, T_1_ (∼12.5 cell doublings) and T_2_ (∼25 doublings). Resultant screen data was analyzed with MAGeCK^61^, demonstrating robust depletion of sgRNAs targeting the promoters of genes selected for proliferation-essentiality and cancer-type susceptibility status, confirming screen efficacy (**Methods, Supplementary Tables 6-7**). We next used CRISPR-SURF^62^ to deconvolve signals from 5p15.33-tiled sgRNAs across datapoints into 2,718 proliferation-essential elements at FDR ≤ 0.05 (median length 470 bp; **Supplementary Table 8**), including strong signals intersecting the *TERT* and *CLPTM1L* promoters. This nominated a large fraction of the locus as proliferation-essential (median ∼40–50% per cell line per timepoint; **Supplementary Table 9**), whereas both melanoma lines showed substantially reduced sensitivity (6.1–17.6% of the locus across timepoints).

To nominate CCVs with the strongest functional support across the four cancers, we integrated MPRA hits with CRISPRi screening data stratified by pleiotropic ASSET signals^5^ **(Fig. 2)**. We defined CCVs as *functional variants* for a given cancer type upon concurrent observation of (i) MPRA-hit status and (ii) colocalization with a CRISPR-SURF-identified regulatory element (FDR ≤ 0.05) in at least one representative cell line for a given cancer type (**Fig. 2**). We then defined variants as *multi-cancer functional variants (MCFVs)* upon annotation as functional variants in more than one cancer type, nominating eight MCFVs. Stratifying MCFVs by corresponding ASSET signal (pairwise *r*² ≥ 0.6; 1kGP p3 EUR; **Fig. 2**), the rs465498 signal in *CLPTM1L* again accounted for the largest set, yielding four MCFVs: rs421629 (*r*² = 0.98 with rs465498), rs11133729 (*r*² = 0.99), rs31487 (*r*² = 0.97), and rs37005 (*r*² = 0.75). Notably, rs421629, rs11133729, and rs31487 were included in the PDAC SuSiE credible set, and rs31487 additionally mapped to the melanoma credible set (**Fig. 2**). Three MCFVs were detected within intron 2 of *TERT* (rs7705526 signal: rs4449583 (*r*² = 0.78); rs11414507 signal: rs74682426 (*r*² = 0.98) and rs56023411 (*r*² = 0.62); **Fig. 2**). Neither of these MCFVs were captured in SuSiE credible sets but were associated with individual cancer types (rs4449583: *P_melanoma_ =* 2.04×10^−15^*, P_lung_EUR_ =* 1.49×10^−20^*, P_lung_EAS_ =* 9.82×10^−43^; rs56023411: *P_melanoma_ =* 1.27×10^−17^). Another *TERT* intron 2 MCFV, rs62332591 was not correlated to any 2021 ASSET signal^5^) at *r*² ≥ 0.6, but was most highly correlated to signal rs2853677 from the 2014 ASSET analysis^4^(*r*^2^ = 0.49).

Having nominated a panel of MCFVs, we next used SNP-centric individual CRISPRi guides to link functional variants to their effector gene(s) (**Fig. 3a-b**). Guided by consistent antagonistic pleiotropy between PDAC and lung cancer in published ASSET analyses^4,5^, we performed CRISPRi to target the respective regions surrounding MCFVs in representative cell lines (PANC-1 for PDAC; A549 for lung cancer) and assessed effects on *TERT* and *CLPTM1L* expression by TaqMan RT-qPCR analysis (**Fig. 3c**; **Methods**). Targeting MCFVs assigned to both the rs7705526 (blue, **Fig. 3**) and rs11414507 (red, **Fig. 3**) ASSET signals in *TERT*’s intron 2 robustly decreased *TERT* expression in tested cells, with weak or no effects on *CLPTM1L*. By contrast, three MCFVs assigned to the rs465498 signal (purple, **Fig. 3**), comprising CCVs intronic and upstream of *CLPTM1L*, produced more modest and heterogeneous transcriptional effects. Perturbation of the respective regions surrounding MCFVs rs31487 and rs37005 reduced *TERT* expression (with rs37005 also decreasing *CLPTM1L* in A549 cells), whereas perturbation of the region containing rs421629 (within *CLPTM1L* intron 16) yielded opposing *TERT* effects between cell types (increased in PANC-1 but decreased in A549 for a subset of sgRNAs) (**Fig. 3c**).

**Fig. 3:**
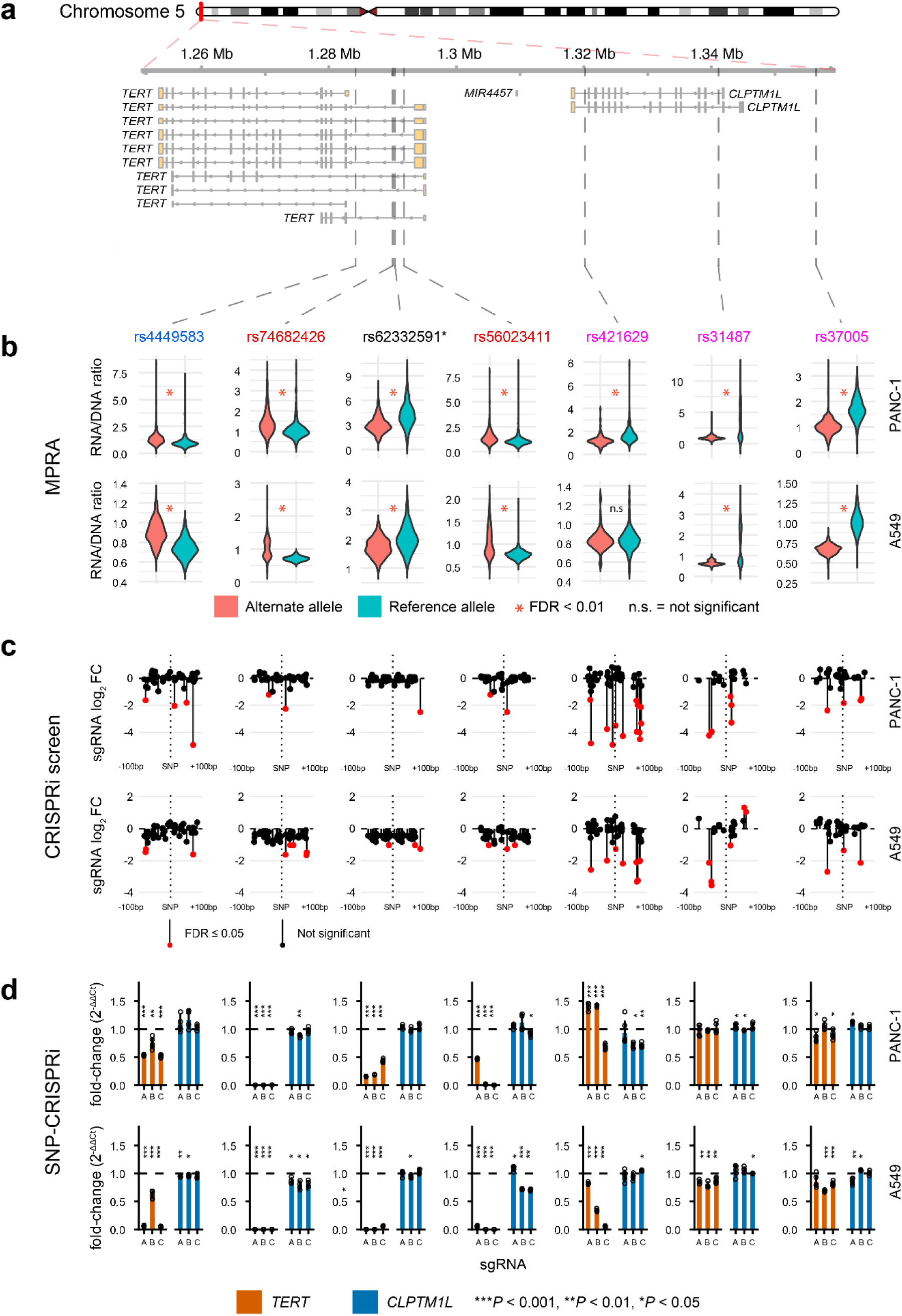
MCFV-targeted CRISPRi nominates TERT and CLPTM1L as effector genes. **a,** We identified eight MCFVs (rs11133729 assayed as part of subsequent VNTR-focused analysis) across the four cancer types as MCFVs corresponding to three 2021 ASSET signals (LD *r*^2^ ≥ 0.6 EUR to ASSET signal lead SNP rs7705526 in purple, rs35226131 in yellow and rs465498 in red, MCFV rs62332591 did not meet LD correlation threshold to ASSET signal-tagging SNP**\***). We chose to highlight screening results and perform subsequent experiments for seven MCFVs in PANC-1 PDAC cells and A549 lung cancer cells due to antagonistic pleiotropy across signals between these two cancer types. **b,** MPRA results for MCFVs revealed significant allelic effects on transcriptional output. Significance indicators reflect P-Value derived from linear model comparing aggregate normalized RNA/DNA TPM ratios from 35 unique tag sequences assigned to both reference (blue) and alternate alleles (red). **c,** While our CRISPRi screening strategy aimed to nominate proliferation-essential elements agnostic to the location of 5p15.33 CCVs, CCV-focused analysis of MCFV-harboring loci (± 100 bp centered on each CCV) revealed significant depletion of sgRNAs targeting respective loci in PANC-1 and A549 cells (red dots indicate FDR ≥ 0.05 depletion of sgRNA with y-axis showing log_2_ fold change as determined by CRISPR-SURF following 25 cumulative screen doublings). **d,** MCFV-targeted CRISPRi using three distinct sgRNAs (x-axis; sgRNAs A, B and C) reduces *TERT* and *CLPTM1L* mRNA expression relative to a non-targeting control sgRNA (y-axis; experimental sgRNA 2^−ΔΔCt^ normalized to 2^−ΔΔCt^ of non-targeting sgRNA, * indicates significance under two-sided t-test). Variants attributable to the rs465498 ASSET signal across *CLPTM1L* demonstrated more modest effects relative to *TERT*, with rs421629 showing a striking effect on *TERT* expression across cell lines, with significant upregulation observed in PANC-1 cells upon epigenetic silencing contrasted to downregulation when using the same sgRNAs in A549, suggesting the presence of a context-dependent regulatory element. Color of text of SNPs above panel A indicates the signal each SNP is part of (in accordance with color scheme in Fig. 2)

### A variable number tandem repeat (VNTR) polymorphism in *CLPTM1L* is part of the signal in *CLPTM1L* tagged by rs465498

SNP rs11133729 was identified as a MCFV mapping to the ASSET signal tagged by rs465498 (*r*^2^ = 0.99, **Fig. 4a**). Located in intron 9 of *CLPTM1L,* this region was annotated by prominent enhancer/promoter marks across ENCODE cell lines (**Fig. 4a)** and was nominated as proliferation-essential in 7/8 cell lines (**Fig. 4b**). This SNP maps within the bounds of a variable number tandem repeat (VNTR) (2,728 bp: chr5:1,326,295-1,329,022, GRCh38, denoted VNTR1), itself located 371 bp upstream of a second VNTR (669 bp, denoted VNTR2, chr5:1,329,508-1,330,176, GRCh38), with both VNTRs comprising distinct 29 bp consensus repeat units (RU) (**Fig. 4c).** Given emergent appreciation of repeat polymorphisms as underlying drivers of GWAS association signals^41,63–65^, we next aimed to determine whether *CLPMT1L* intron 9 VNTR polymorphisms might represent CCVs at 5p15.33. To achieve this, we utilized a joint genotyping strategy to infer read-phased VNTR and SNP haplotypes from PacBio amplicon reads within *CLPTM1L* intron 9 across 484 1kGP EUR samples as well as DNA samples from 940 patients diagnosed with PDAC and 870 controls from the PanScan I^66^ and II^67^ GWAS phases (see **Methods, Supplementary Tables 10-12**). In the PDAC GWAS cohort, we resolved 459 distinct VNTR1 alleles spanning lengths of 2,149–4,560 bp, with a broadly trimodal frequency distribution (peaks at 2,586 bp, AF = 0.07; 2,789 bp, AF = 0.31; 2,934 bp, AF = 0.31; **Fig. 4d**), and VNTR2 alleles spanning 527–1,830 bp with a bimodal distribution (peaks at 730 bp, AF = 0.42; 933 bp, AF = 0.36; **Fig. 4d**).

**Fig. 4:**
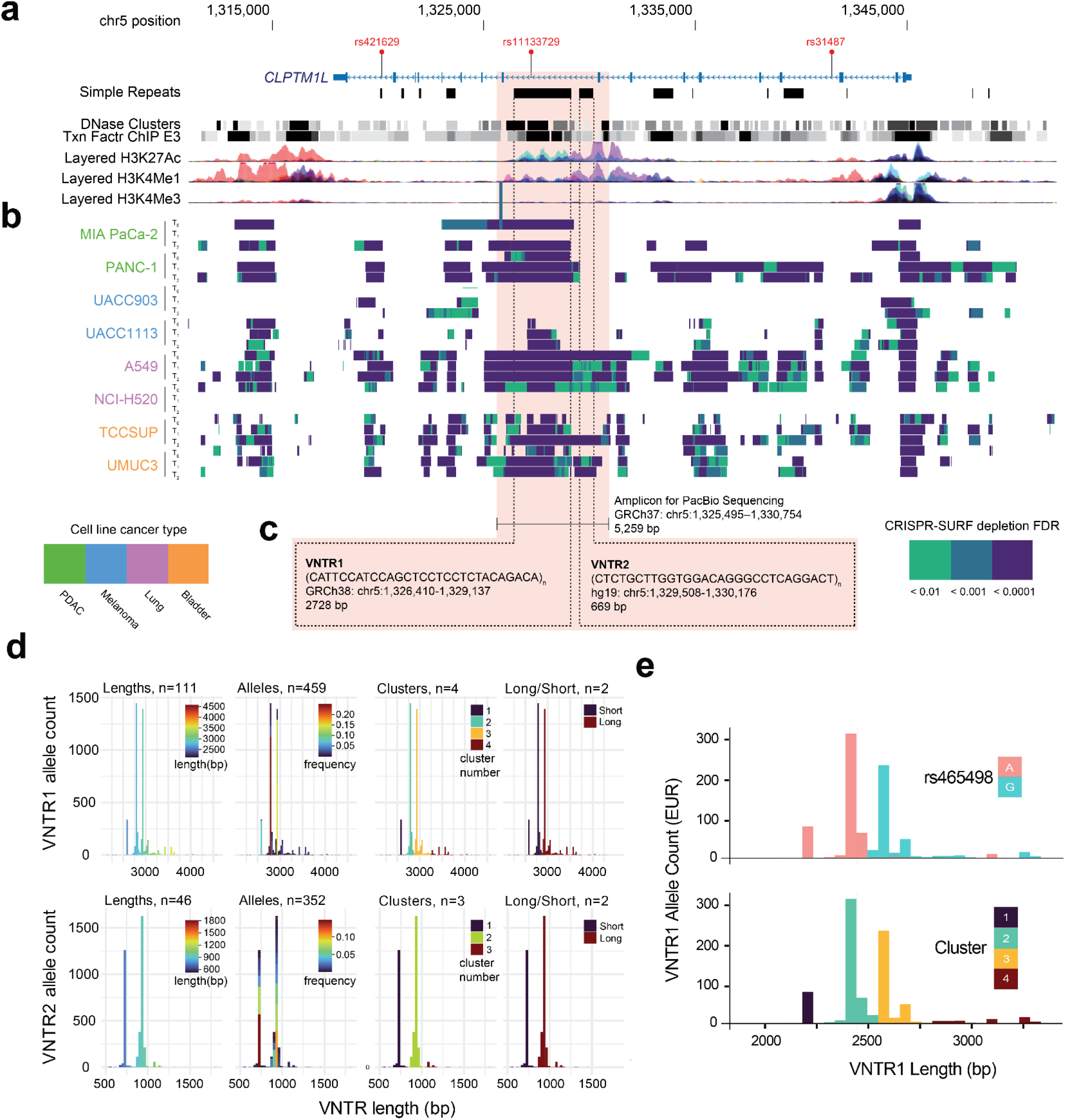
Tiled CRISPRi proliferation screen nominated VNTRs represent novel CCVs for PDAC. **a,** Functional screening of the 5p15.33 locus nominated three MCFVs intronic to *CLPTM1L*. One such MCFV, rs11133729, resides in intron nine of *CLPTM1L*, a region enriched for enhancer-associated histone marks, DNAse hypersensitivity marks and transcription factor ChIP-Seq peaks across seven ENCODE cell lines as demonstrated from the UCSC genome browser’s ENCODE regulation track (GRCh37). **b**, Analysis of tiled CRISPRi screening data of the 5p15.33 locus using CRISPR-SURF nominated a considerable proportion of *CLPTM1L* as proliferation-essential, including prominent depletion of sgRNAs within intron 9. Aligned tracks display proliferation-essential elements from CRISPR-SURF colored by magnitude of CRISPR-SURF depletion FDR. For each cell line (colored by representative cancer type), screening data was analyzed across three timepoints (T_0_: immediately following puromycin selection and recovery, T_1_: ∼ 12.5 cumulative screen doublings, T_2_: ∼ 25 cumulative doublings).**c,** Intron 9 is notable for the presence of two variable number tandem repeat (VNTR) elements. CRISPRi screening across tested cell lines nominated both VNTRs as proliferation-essential elements. As VNTRs are known to be highly polymorphic, we utilized PacBio long-read amplicon sequencing to genotype both VNTRs in 1kGP and PDAC GWAS samples. **d,** VNTR genotyping in 1,810 PDAC cases and controls revealed trimodal (VNTR1) and bimodal (VNTR2) allele length distributions. (VNTR allele count plotted on y-axis with corresponding VNTR length on x-axis) Bar plots colored as follows; aggregate VNTR lengths colored by corresponding length, distinct alleles by observed frequency, clusters by k-means cluster index, long/short by binary indicator). **e,** Read-based phasing of VNTR and SNP alleles in 484 1kGP EUR samples demonstrated co-segregation of the rs465498 PDAC risk haplotype (top panel) with longer VNTR1 alleles (clusters, bottom panel).

Genetic association tests for VNTRs have often grouped alleles as long vs. short to overcome the challenges in genotyping with traditional sequencing and PCR-based methods^41,44,68^. Given that TRGT outputs full VNTR sequences as alleles, we assessed VNTR associations with individual risk signals by encoding VNTR genotypes from the 1,810 PanScan samples in four different ways, with increasing levels of aggregation, as follows (**Fig. 4d**); (i) discrete VNTR alleles (each unique haploid VNTR sequence as its own allele, VNTR1_allele_n, VNTR2_allele_n, both represented by allele length followed by unique index number (n)), (ii) aggregate VNTR lengths (collapsing alleles of identical length regardless of sequence, VNTR1_length, VNTR2_length), (iii) data-driven length classes defined by applying k-means clustering to observed allele-length distributions (k selected based on visual inspection of allele-length distributions, VNTR1_cluster, VNTR2_cluster, yielding k = 4 for VNTR1 (cluster 1: 2,149–2,673 bp; cluster 2: 2,702–2,872 bp; cluster 3: 2,876–3,224 bp; cluster 4: 3,253–4,560 bp) and k = 3 for VNTR2 (cluster 1: 527–817 bp; cluster 2: 846–1,020 bp; cluster 3: 1,049–1,803 bp) and (iv) a binary long/short (VNTR1_LS, VNTR2_LS) encoding obtained by collapsing adjacent length classes within each VNTR (VNTR1: short = clusters 1–2, long = clusters 3–4; VNTR2: short = clusters 1–2, long = clusters 2-3)). We found VNTR1_LS long encodings were linked to the PDAC risk haplotype marked by the rs465498 ASSET signal (rs31490-A, **Fig. 4e**; pairwise LD between VNTR alleles and rs465498; VNTR1 long: *r*² = 0.83, D′ = 0.99; VNTR2 long: *r*² = 0.70, D′ = 0.99), which motivated subsequent analyses to determine whether VNTR alleles comprise credible causal variants for this signal.

### VNTR alleles associate with PDAC risk

PacBio-derived VNTR alleles were first tested for association with PDAC risk in 1,810 directly sequenced PanScan I and II DNA samples (940 PDAC and 870 controls, see **Methods**). Multiple VNTR1 encodings were associated with PDAC (**Supplementary Table 13**). Among specific VNTR1 alleles, a VNTR1 2,934 bp allele (2934_30) exhibited the most significant association (VNTR1_allele 2934_30; AF = 0.14, OR = 1.52, *P* = 3.25×10^−5^) when compared to all other alleles. Aggregate VNTR1 long alleles also demonstrated significant association with PDAC (VNTR1_LS long; AF = 0.52, OR = 1.22, *P* = 2.54×10^−3^), with VNTR1-cluster 3 demonstrating comparable results (VNTR1_cluster 3; AF = 0.44, OR = 1.20, *P* = 7.73×10^−3^). By contrast, both common and infrequent VNTR2 allele encodings yielded only nominally significant associations, including VNTR2-long (VNTR2_LS long; AF = 0.55, OR = 1.15, *P* = 0.05) and a VNTR2 962-bp allele (VNTR2_allele 962; AF = 0.006, OR = 1.50, *P* = 0.01).

To increase statistical power, we imputed VNTR genotypes into a larger PDAC GWAS dataset. Using PacBio-derived VNTR encodings and SNP calls from the 1,810 PanScan I & II samples described above, we imputed each of the four VNTR genotype encodings (**Fig. 4d**) as distinct multiallelic variants into the remaining samples from the PanScan/PanC4 PDAC meta-GWAS^66,67^ (including an additional 8,066 cases and 11,208 controls; **Methods**). After imputation, we filtered VNTR and SNP alleles (MAF ≥ 0.01, imputation quality score R2 ≥ 0.3) prior to application of the same logistic model applied to directly genotyped samples and subsequent meta-analysis of each cohort. In agreement with the original GWAS, SNP rs31490 exhibited the most significant association^46^ (**Fig. 5b**; OR = 1.20, *P* = 2.33×10^−16^). Results for imputed VNTR alleles largely replicated the analysis of directly genotyped samples **(Supplementary Table 14)**. Of the four VNTR encodings, VNTR1 cluster 3 emerged as the most significant VNTR association (OR = 1.20, *P* = 8.86×10^−15^). We also replicated significant effects for long/short VNTR1 alleles (OR = 1.20, *P* = 4.00×10^−15^). Of the discrete VNTR1 alleles, the 2,934 bp sequence (2934_30) which previously comprised the most significant VNTR association in the pilot analysis retained genome-wide significance (OR = 1.25, *P* = 1.19×10^−9^). Collective VNTR2 lengths of 933 bp were the only significant (at *P* ≤ 5×10^−8^) VNTR2 encoding (OR = 1.17, *P* = 5.36×10^−11^).

**Fig. 5:**
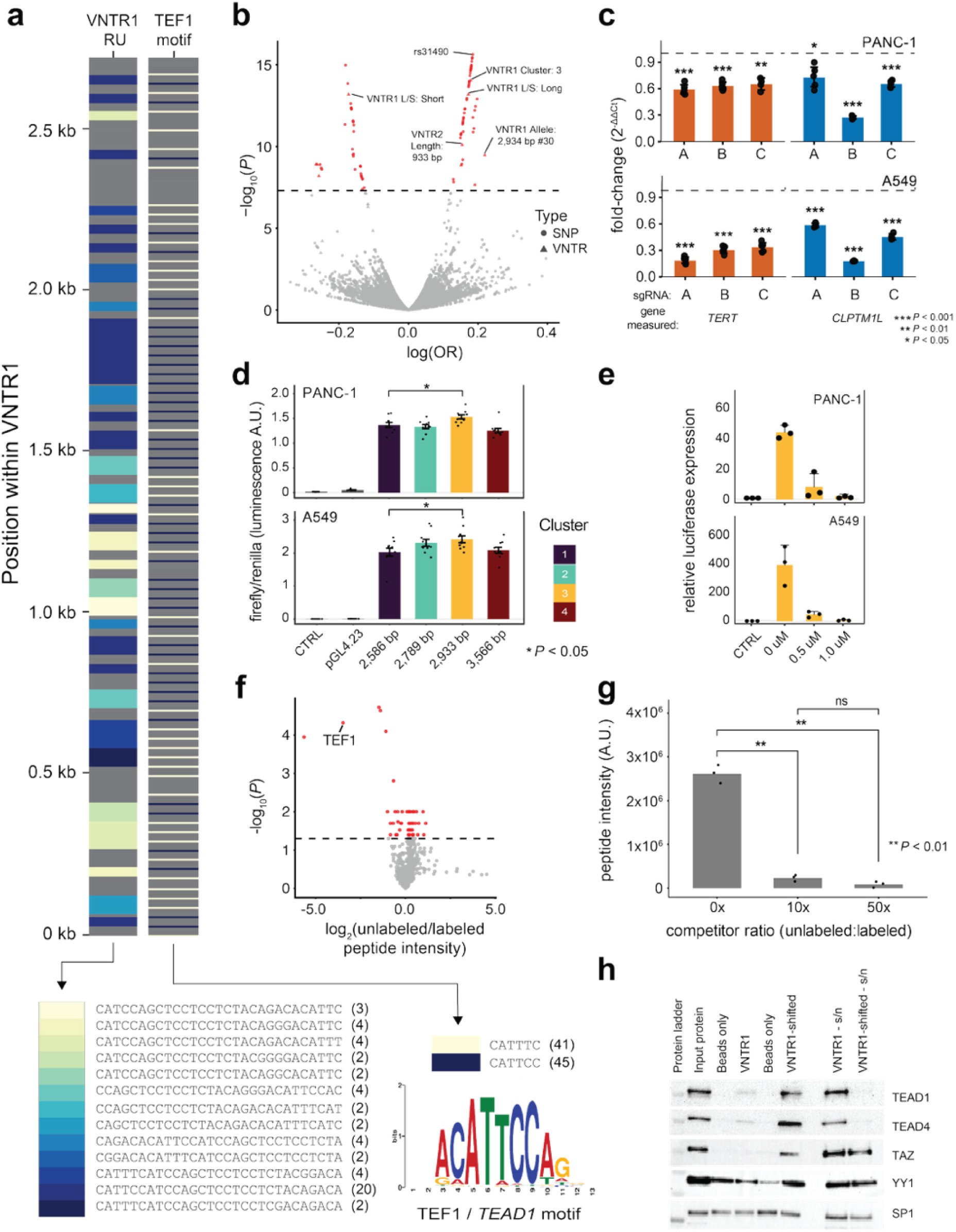
VNTR1 is a Hippo-dependent regulator of TERT and CLPTM1L. **a,** Motifscope plot of VNTR1 as comprised of tandemly repeating 29bp repeats (most frequently observed repeat units are color-coded and shown at bottom left) which themselves are enriched for TEF1 binding sites (canonical TEF1 binding motifs shown in blue and cream, bottom right). **b,** Imputation of VNTR alleles into full GWAS (9,040 PDACs, 12,496 controls) revealed strong association for VNTR alleles (y-axis; −log10 *P*, red: *P* ≤ 5×10⁻⁸; logistic regression of SNP/VNTR dosage adjusted for age, sex sample site and principle components) **c,** VNTR1-targeted CRISPRi using three distinct sgRNAs (x-axis; sgRNAs A, B and C) reduces *TERT* and *CLPTM1L* expression (y-axis; experimental sgRNA 2^−ΔΔCt^ normalized to 2^−ΔΔCt^ of non-targeting sgRNA, * indicates significance under two-sided t-test) **d,** Dual luciferase reporter assays incorporating cluster-representative VNTR1 sequences demonstrated strong VNTR1 enhancer activity in PANC-1 and A549 cells; cluster 3 alleles show significantly higher luciferase activity as compared to cluster 1 (Welch’s t-test, raw firefly/renilla shown due to intensity of signal (**Methods**)). **e,** Small molecule inhibition of TEF1/YAP binding negates VNTR1 enhancer activity, indicating Hippo pathway dependence (y-axis; firefly/renilla luminescence normalized to negative control plasmid, **Methods**). **f,** DNA pulldown/mass spectrometry comparing peptide intensity following incubation of Mia PaCa-2 nuclear lysate with biotinylated DNA probes containing consensus VNTR1 repeat unit and addition of 50-fold excess untagged competitor probe reveals VNTR1-specific TEF1 binding (x-axis; log_2_ unlabeled/labelled peptide intensity, y-axis: −log_10_ *P,* one-sided *t*-test); **g**, Bar plot of TEF1-specific specific mass-spectrometry results, 10 and 50-fold excess unlabeled competitor negate TEF1 intensity (independent-samples t-test). **h,** Western blot following DNA oligo pulldown experiments confirms VNTR1-specific TEF1 binding. Binding is only seen for VNTR1-shifted probe (sequence of RU offset by 15 bp) to accommodate TF motif in RU junction. Similar binding was observed for Hippo pathway factors TEF-3 (gene name: *TEAD4*) and WW domain-containing transcription regulator protein 1 (gene name: *WWTR1*; also named *TAZ*). Nonspecific binding to beads and probes was seen for YY1 and SP1. “s/n” indicates flowthrough protein levels after the pulldown

Conditioning the association analysis on VNTR1 cluster 3 by including corresponding VNTR encoding dosages as a fixed covariate resulted in marked attenuation of the signal for lead SNP rs31490 (*P_VNTR1_cluster3_* = 9.30×10^−3^). Conditioning on VNTR2 lengths of 933 bp resulted in more modest attenuation of the rs31490 signal (*P*_VNTR2_933bp_ = 4.13×10^−6^). Conditioning the analysis on rs31490 likewise attenuated the VNTR1 cluster 3 association (*P_rs31490_* = 0.18). Alternatively, conditioning on SNPs known to tag PDAC signals confirmed linkage with the rs31490 haplotype (*P*-values for VNTR1 cluster 3 conditioned on PDAC signal SNPs: *P*_conditional_rs451360_ = 1.37×10^−5^, *P*_conditional_rs2735940_ = 3.62×10^−9^, *P*_conditional_rs2853677_ = 2.79×10^−7^, **Supplementary Table 14**).

### VNTR1 is a potent enhancer at the 5p15.33 locus

Given strong evidence that VNTR1 polymorphisms comprise plausible CCVs as part of the rs31490/rs465498-tagging haplotype (**Fig. 4d**), we next sought to characterize the biological mechanism underlying VNTR1-enhancer activity. *In*-*silico* analysis of VNTR1-transcription factor binding sites using TRAP^69^ revealed significant enrichment for TEA Domain Family Member 1 (TEF-1) motifs (*P_nominal_* < 5.0×10^−4^, **Fig. 5a**, **Supplementary Table 15**). TEF-1, encoded by *TEAD1,* is a key transcription factor member of the Hippo signaling pathway involved in regulation of apoptosis and proliferation^70–73^.

Similar to our approach for small sequence variants, we individually assessed VNTR1 using CRISPRi and individual luciferase reporter assays to assess functionality. CRISPRi repression of VNTR1 reduced expression of both *TERT* and *CLPTM1L* relative to negative control sgRNA in PANC-1 and A549 cells (**Fig. 5c**). VNTR1-sgRNA-A, which directly targets the region harboring rs11133729 in VNTR1, reduced *TERT* (fold-change = 0.19, *P* = 6.69×10⁻⁵) and *CLPTM1L* (fold-change = 0.58, *P* = 1.30×10⁻⁴) expression in A549 cells, and similarly decreased *TERT* (fold-change = 0.59, *P* = 2.78×10⁻⁴) and *CLPTM1L* (fold-change = 0.72, *P* = 1.43×10⁻^2^) expression in PANC-1 cells. Luciferase reporter assays of VNTR1 alleles representative of each of the cluster-genotype encodings confirmed potent enhancer activity in both cell lines (median fold change vs. negative control: PANC-1 = 74; A549 = 609; **Fig. 5d**). Enhancer strength varied across constructs representative of VNTR1 cluster encodings: relative to the shortest tested allele (2,586 bp; assigned to cluster 1), a 2,933 bp sequence assigned to VNTR1 cluster 3 demonstrated higher activity in PANC-1 (fold-change = 1.28, *P* = 2.69×10⁻⁴) and A549 (fold-change = 1.20, *P* =2.23×10⁻²) cells.

Having established VNTR1 as a potent enhancer for *TERT* and *CLPTM1L*, we proceeded to characterize the transcription factors mediating this effect. We performed DNA pulldown experiments coupled with mass spectrometry by incubating biotinylated VNTR1 repeat units with MIA PaCa-2 nuclear lysate. This analysis identified robust binding of TEF-1 peptides to VNTR1 sequences (**Fig. 5f, Supplementary Table 16**). Notably, competitor experiments led to a striking depletion of TEF-1 (**Fig. 5g**), indicating highly sequence-specific protein-DNA interactions. This finding was validated by oligo pulldown experiments followed by western blot showing robust binding of TEF-1 (*TEAD1*) as well as additional Hippo pathway family members TEF-3 (*TEAD4*) and WW domain-containing transcription regulator protein 1 (*WWTR1*; also named *TAZ*) to the VNTR1 repeat motif (**Fig. 5h**). Finally, we sought to determine to what degree VNTR1-luciferase activity was attributable to Hippo-pathway transcription factor (TF) signaling. Disruption of TEF-YAP complexes with verteporfin, a small molecule inhibitor of TEF-YAP binding^74,75^, markedly reduced VNTR1-driven reporter activity across constructs in both cell lines (**Fig. 5e**), indicating that VNTR1 enhancer function depends on Hippo-pathway transcriptional complexes. Collectively, these data support VNTR1 polymorphisms as MCFVs at 5p15.33 with regulatory implications for both *TERT* and *CLPTM1L*.

## Discussion

Here, we provide a comprehensive dissection of functional *cis-*regulatory CCVs at 5p15.33. By identifying functional variants which mediate causal effects on both target gene expression and cell proliferation, our work furthers understanding of the mechanisms underpinning susceptibility for multiple cancers. We derived an integrated functional map for 116 5p15.33 CCVs and identified eight MCFVs across three distinct GWAS signals, demonstrating allele preferential *cis*-regulatory effects that localized to proliferation-essential regulatory elements across cancer cell lines from four cancer types. Additionally, we build on findings from transcriptome-wide association studies^34^ and post-GWAS functional analyses which support the role of *CLPTM1L* as a possible target gene for the GWAS signals at 5p15.33.

Antagonistic pleiotropy between cancers has long been a confounding factor at 5p15.33. Context-dependent telomerase biology has been hypothesized as a possible mediator of such effects, although direct analysis of telomere length across cancer types has yielded inconclusive results^76–84^. Our analysis marks a major step towards understanding how these differences may be mediated at a *cis-*regulatory level. For all eight MCFVs we identified, MPRA allelic effects demonstrated consistent directionalities across cancer types, suggesting that differences in phenotypic outcome may largely be driven by effects of downstream target genes and pathways, epigenetic factors, or potentially by interactions with environmental factors. Our tiled CRISPRi screen supported this model, with MPRA-functional CCVs intersecting elements nominated as putative enhancers. Focused interrogation of these variants with CRISPRi proved largely consistent with this model except for rs421629. Our findings suggest that this SNP occupies a possible context-dependent regulatory element whose net effect on *TERT* may switch between repressive and activating effects depending on the cell type. An alternative, non-mutually exclusive possibility is that perturbation of this element reconfigures the local regulatory circuit, such that inhibiting rs421629-dependent activity unmasks or engages distinct enhancers or promoters in a cell type-specific manner. This reversal provides a plausible *cis-*regulatory mechanism for antagonistic pleiotropy at 5p15.33. Motif analysis indicated that the PDAC/melanoma risk allele (rs421629-G) may introduce high affinity binding motifs for NFATC2, raising the possibility that allele-specific TF recruitment, combined with lineage-specific cofactor and chromatin environment, may toggle the element between enhancer- and repressor-like states. While additional work is required, our data highlights how cell type-specific wiring of a single *cis*-regulatory element may contribute to antagonistic pleiotropy at 5p15.33.

Our analysis identified a functional VNTR polymorphism as an additional CCV for the signal at *CLPTM1L* marked by rs465498, where both VNTR and SNP alleles may jointly underpin the observed association. We characterized VNTR1 as a potent enhancer of both *TERT* and *CLPTM1L* with activity driven by Hippo pathway TFs. Notably given antagonistic pleiotropy for this signal, Hippo pathway factors have been implicated in both oncogenic and tumor suppressive mechanisms across cancer types^85^. We also note four MCFVs in intron 2 of *TERT* intersecting repeat elements, corresponding to several independent association signals. Our analysis supports recent studies which posit VNTRs as a considerable source of missing heritability^86,87^. While post-GWAS analyses are largely focused on the identification of causal SNPs and their relationships to target genes, there are several examples of GWAS signals tagging more complex variants (e.g., in *TERT*^41^, *CTRB2*^88^, *EGR2*^65^). Repeat elements are pervasive throughout the genome, with a substantial body of evidence supporting their role as *cis*-regulatory elements^89–91^. Recent studies have estimated VNTR polymorphisms to underly prominent glaucoma and colorectal cancer GWAS signals^63^. Moreover, studies have estimated 11.4% of structural variants and simple tandem repeats to be in LD with SNPs associated with 701 GWAS traits^86^. Identification of cryptic functional alleles is particularly pertinent at complex loci such as 5p15.33, where multiple independent signals and context-dependent regulatory elements confound deeper understanding of cancer susceptibility. To accelerate functional characterization at such loci, it is imperative that repeat polymorphisms are included into GWAS imputation reference panels given recent advances in long-read sequencing technologies.

While this work provides a comprehensive map of cross-cancer regulatory elements at 5p15.33, our approach is not without limitations. While this locus has been implicated in numerous common cancers, we specifically fine-mapped and functionally interrogated four representative cancer types. While we assessed 11 pleiotropic signals, functional screening of only four cancer types may have limited our sensitivity to identify MCFVs for some signals. Furthermore, all experiments were performed in cancer cell lines, which do not fully recapitulate the physiological and epigenetic context in which germline variants act. Future studies should aim to investigate the effects of MCFVs in primary cells and 3D cell culture models to better understand the mechanisms of pleiotropy at 5p15.33. In addition, lung cancer GWAS across ancestries have shown that the signal tagged by rs2736100 is specific to lung adenocarcinoma (EAS), whereas the signal tagged by rs6554758 is more strongly associated with squamous cell carcinoma^49^. MCFVs assigned to each signal were supported by evidence from both adenocarcinoma (A549) and squamous cell carcinoma (NCI-H520) models; however, subsequent studies should examine how these functional alleles act across lung cancer subtypes. More broadly, future work should also endeavor to test functional alleles in contexts that model both genetic and environmental stressors. Because MCFV and VNTR characterization assays were conducted in *TERT* promoter wild type PANC-1 and A549 cells^7,92^, it would also be appropriate to assay the effect of these polymorphisms in the context of somatically-mutated *TERT* promoters, such as in UACC903 or UACC1113 melanoma cell lines. Lastly, our CRISPRi screens interrogated regulatory element function without directly resolving allelic effects; lenti-MPRA or CRISPR base-editing approaches could quantify allele-specific transcriptional consequences within native chromatin contexts in models not amenable to transient transfection.

In summary, our model identifies multiple potential causal variants across the region, including several linked to the same signals. In addition, this work implicates rs421629 as part of a regulatory element with seemingly dual functions on target gene expression, highlighting *cis*-regulatory mechanisms as a potential mediatory of antagonistic pleiotropy at 5p15. Finally, we demonstrate that VNTR polymorphisms contribute to the *CLPTM1L* signal marked by rs465498, suggesting contributions from both simple and complex polymorphisms, as well as demonstrating a potentially broader role of repeat polymorphism at complex susceptibility loci.

## Data Availability

All data produced in the present study are available upon reasonable request to the authors

---

**Supplementary Fig. 1:**
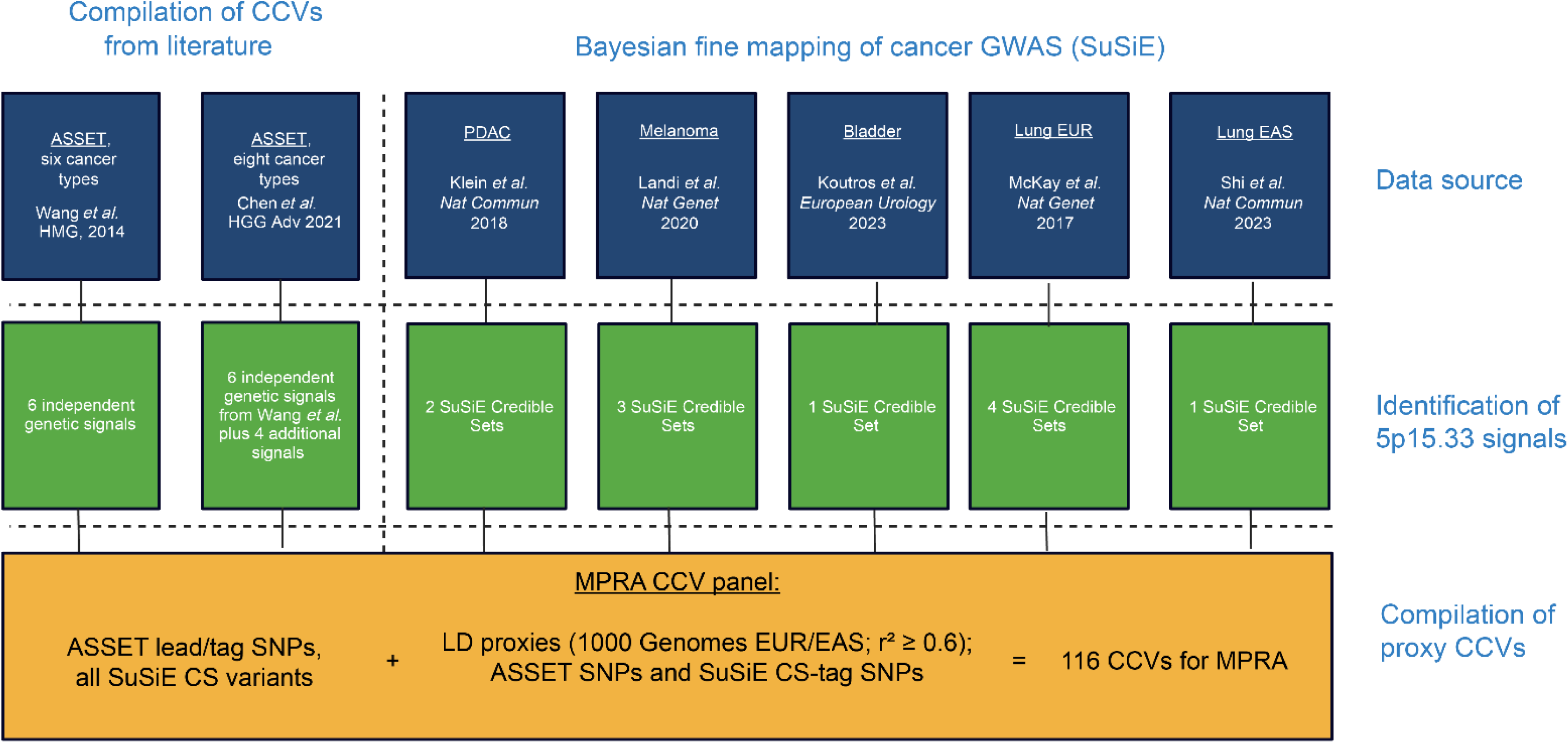
Credible causal variant (CCV) inclusion criteria for MPRA testing. Two studies have comprehensively identified common risk variants at 5p15.33, which are associated with the risk of several cancer types. Wang et al. in 2014 used the Association analysis based on SubSETs (ASSET) method of conditional and joint meta-analysis of GWAS summary statistics to identify six distinct signals at the locus, which was expanded upon by Chen et al. in 2021, identifying an additional four independent signals. These papers served as a starting point for CCV compilation. These signals were orthogonally supplemented by the application of the Bayesian fine-mapping tool Sum of Single Effects (SuSiE) to individually identify independent signals from GWAS summary statistics for PDAC, lung cancer, bladder cancer and melanoma. GWAS for lung adenocarcinoma were separately analyzed for European and East Asian ancestries. We supplemented ASSET-derived variants and variants assigned to credible sets by identifying signal-tagging ASSET variants along with those variants with the highest PIP for a given credible set and subsequently including correlated proxies with LD r^2^≥0.6 to derive a final panel of 116 CCVs for inclusion in the MPRA pool.

**Supplementary Fig. 2:**
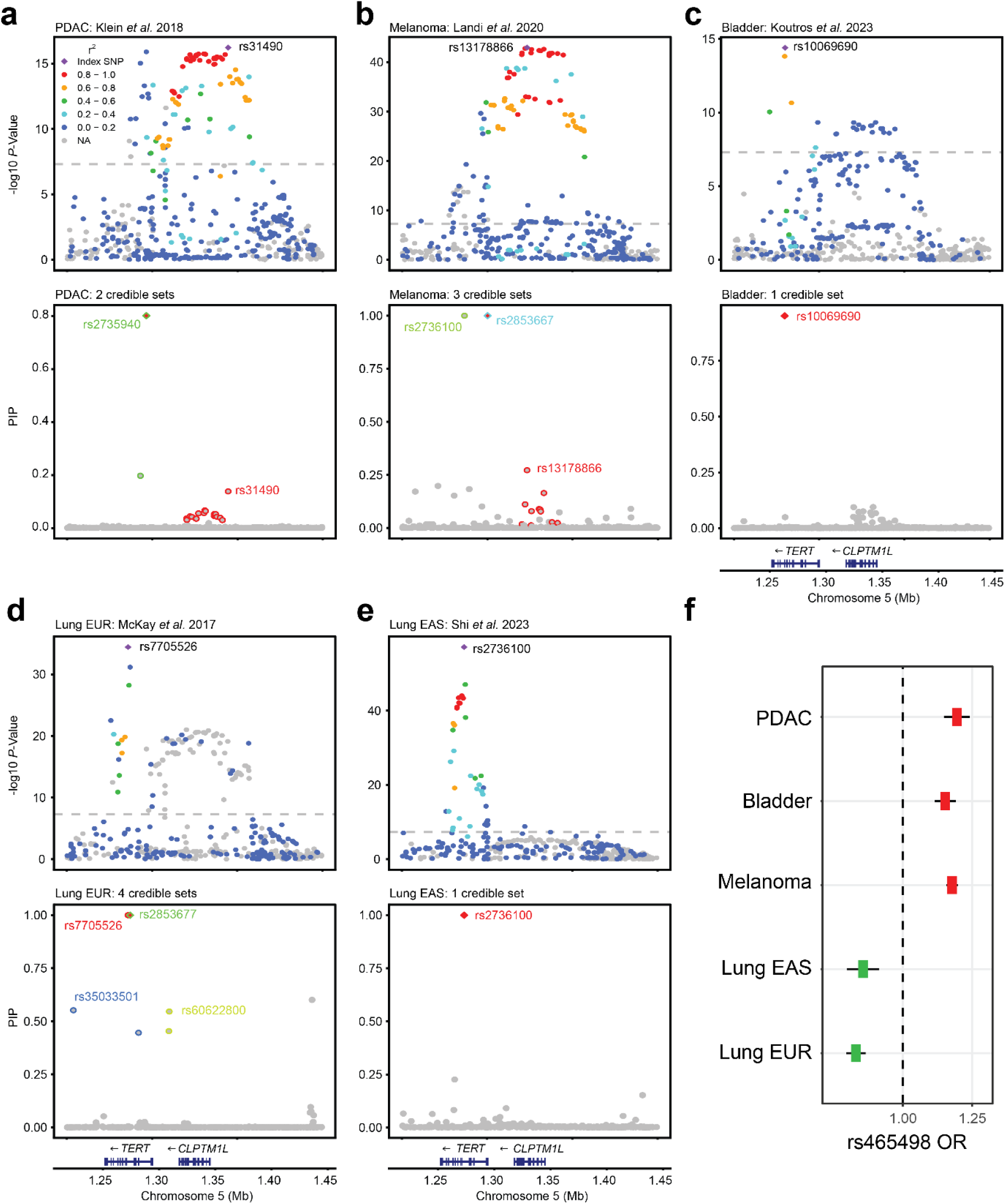
Fine-mapping of 5p15.33 implicated cancer GWAS. GWAS signals at the 5p15.33 locus in fine mapped cancers with resultant SuSiE credible sets are shown with posterior inclusion probabilities (PIPs) for each variant shown below for; **a,** PDAC, **b,** melanoma, **c,** bladder cancer, **d,** European (EUR) lung cancer, and **e,** East-Asian (EAS) lung cancer. **f,** Odds ratios and 95% confidence intervals across all fine-mapped cancers, showing antagonistic pleiotropy between PDAC, melanoma, bladder cancer and lung cancer EUR/EAS GWAS for one of the signals at chr5p15.33 (ASSET signal marked by rs465498).

**Supplementary Fig. 3:**
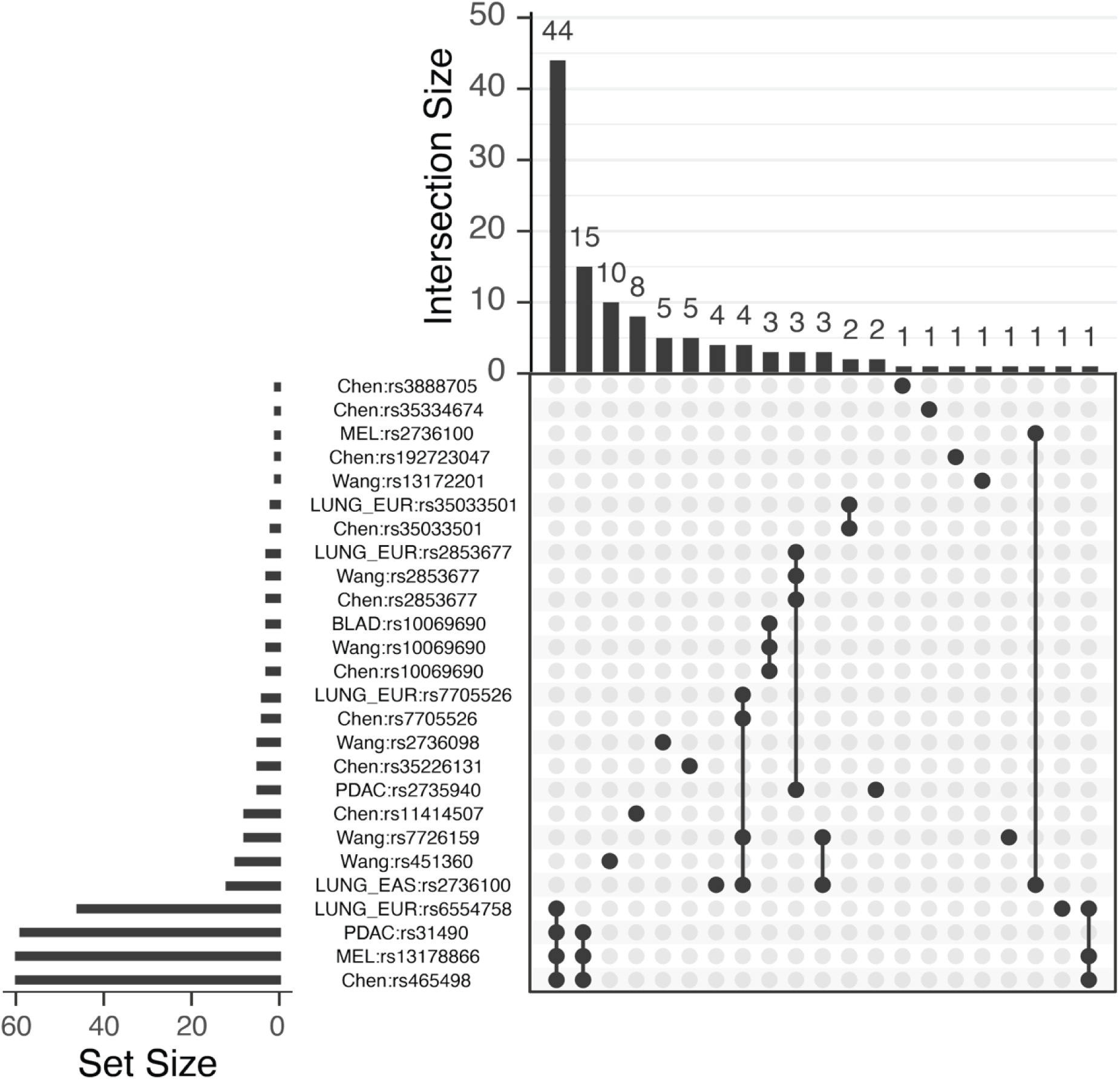
Cross-correlation of ASSET and SuSiE-nominated 5p15.33 signals. UpSet plot showing intersections between SuSiE fine-mapping credible sets and ASSET signal-tagging SNPs identified across five cancer GWAS. Variants were grouped based on linkage disequilibrium (LD r² ≥ 0.6 1kGP EUR). Horizontal bars (left) indicate the total number of variants in each credible set or reported signal. Vertical bars (top) show the size of intersections between sets, with connected dots (matrix below) indicating which credible sets share variants. The most common intersection (n=44 variants) represents a core set of variants shared across multiple cancer types. Cancer type abbreviations: PDAC, pancreatic ductal adenocarcinoma; MEL, melanoma; BLAD, bladder cancer; LUNG_EUR, lung cancer in European ancestry; LUNG_EAS, lung cancer in East Asian ancestry; Chen (Chen et al. ASSET 2021); Wang (Wang et al. ASSET 2014). Results demonstrate extensive sharing of credible causal variants across cancer types, with some cancer-specific signals, consistent with pleiotropic regulatory mechanisms at this locus.

**Supplementary Fig. 4:**
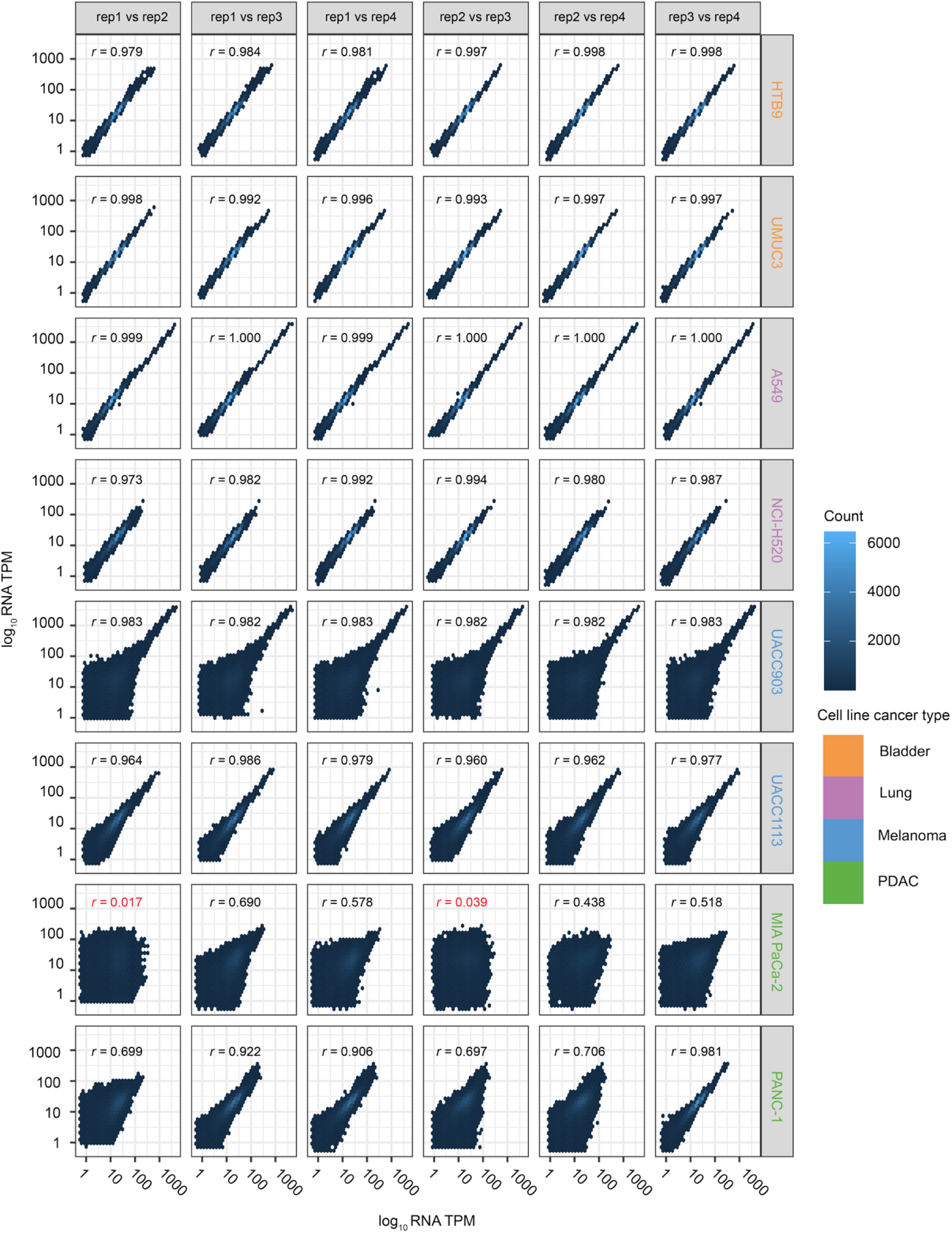
Inter-replicate correlation of MPRA experiments. Pairwise correlation matrices showing inter-replicate concordance for MPRA expression measurements (log_10_ RNA TPM vs. log_10_ RNA TPM) in four biological replicates across eight cancer cell lines. Each panel displays a hexbin scatter plot with Pearson correlation coefficient (r) for comparisons between replicates (rep1 - rep4). Cell lines tested: 5637 (bladder cancer), UMUC3 (bladder cancer), A549 (lung adenocarcinoma), H520 (lung squamous cell carcinoma), UACC903 and UACC1113 (melanoma), MIA PaCa-2 (PDAC), and PANC-1 (PDAC). Point density is indicated by color intensity (blue scale). Most pairwise comparisons showed strong correlations (r > 0.96), except for MIA PaCa-2 replicate 2 (r = 0.017 and r = 0.039, red text) and suggesting technical issues with MIA PaCa-2 replicate 2 which was removed from downstream analyses. High correlations across remaining replicates demonstrate robust and reproducible MPRA measurements for functional variant characterization.

**Supplementary Fig. 5:**
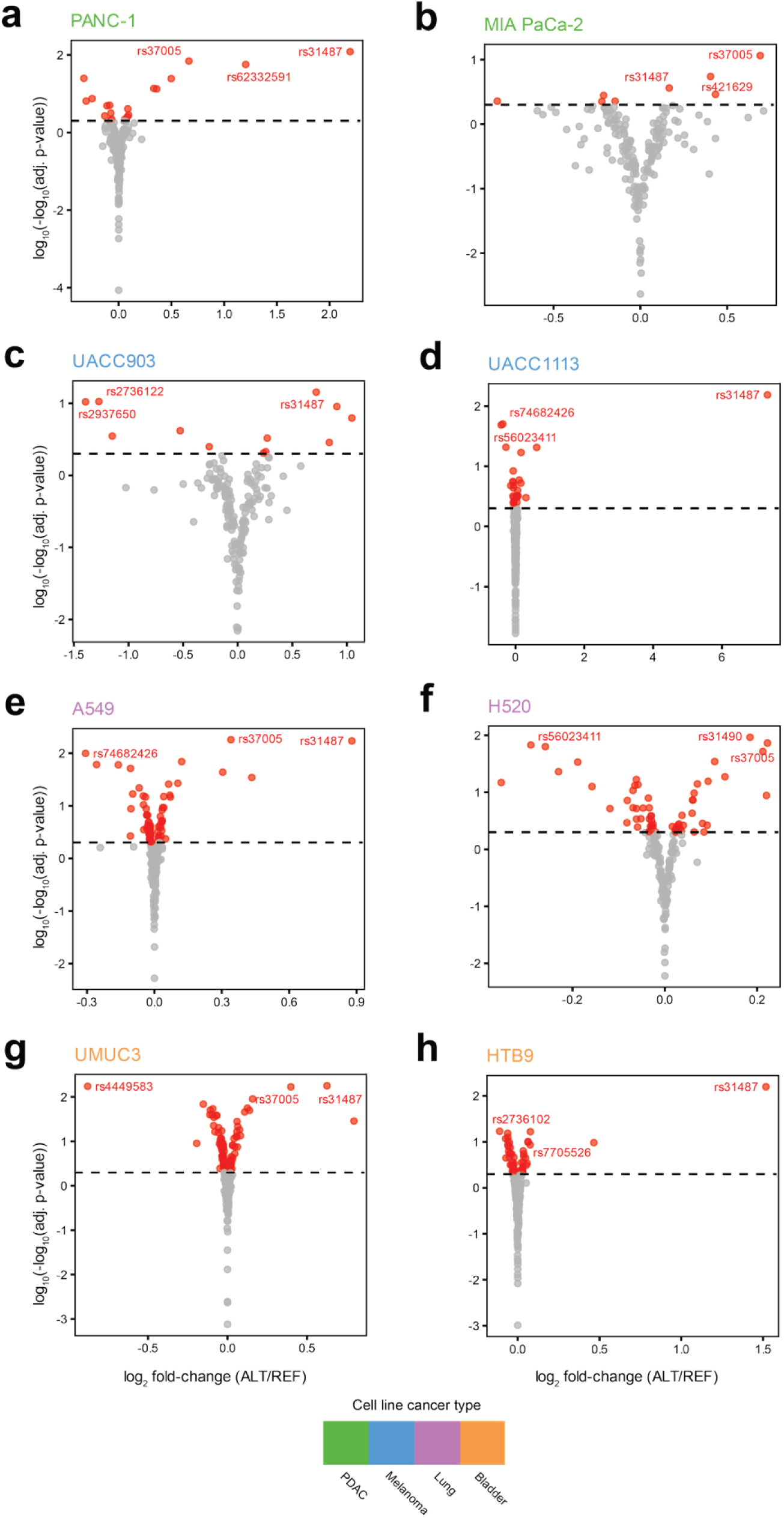
MPRA results for 5p15.33 CCVs revealed variants exhibiting allele preferential activity across eight cell lines. Volcano plots showing allelic effects of MPRA-tested variants in; **a,** PANC-1, **b,** MIA PaCa-2, **c,** UACC903, **d,** UACC1113, **e,** A549, **f,** NCI-H520, **g,** UMUC3, and **h,** 5637 cell lines. X-axis shows log_2_ fold-change in expression between alternative (ALT) and reference (REF) alleles; y-axis shows statistical significance (-log_10_ adjusted p-value). Red points indicate variants with significant allelic activity differences (MPRA-functional variants); gray points and dashed horizontal line indicates non-significant variants (FDR < 0.01). Selected variants of interest are labeled with their rsID numbers. Multiple variants demonstrate significant allelic effects across cancer types, with some showing cell-type-specific regulatory activity. The magnitude and direction of allelic effects vary across cell lines, reflecting tissue-specific regulatory mechanisms.

**Supplementary Fig. 6:**
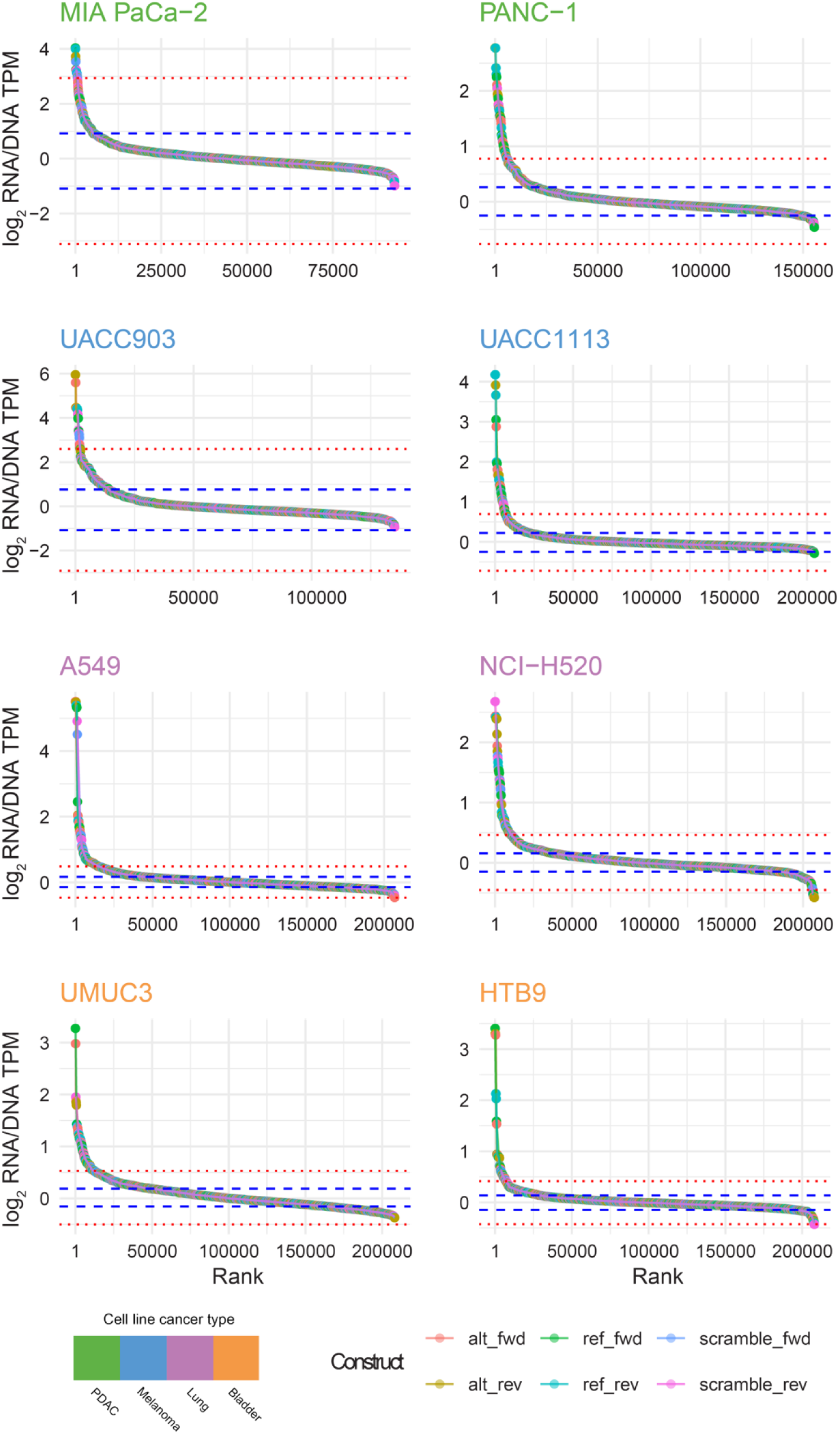
Scrambled control constructs assign MPRA-functional CCVs to active elements. Ranked distribution of log_2_ RNA/DNA TPM values for all tested constructs across eight cancer cell lines: MIA PaCa-2, PANC-1, UACC903, UACC1113, A549, NCI-H520, UMUC3, and 5637/HTB9. Constructs are ranked from highest to lowest expression and colored by type and strand orientation: alternative allele forward strand (alt_fwd, pink), alternative allele reverse strand (alt_rev, orange), reference allele forward strand (ref_fwd, green), reference allele reverse strand (ref_rev, cyan), scrambled control forward strand (scramble_fwd, blue), and scrambled control reverse strand (scramble_rev, purple). Dashed blue lines indicate the first quartile (Q1, lower) and third quartile (Q3, upper) of scrambled control expression within each cell line; dotted red lines indicate Q1-IQR (lower suppressor threshold) and Q3+IQR (upper activator threshold). Alleles with mean expression above Q3 were classified as putative activators; those below Q1 were classified as putative suppressors. CCVs meeting criteria for MPRA-functionality (FDR < 0.01, |fold-change| ≥ 15%, and activator or suppressor classification) demonstrate activity substantially divergent from transcriptionally inert scrambled controls, indicating genuine regulatory function.

## Methods

### Cell culture

UACC903 and UACC1113 melanoma cell lines were obtained from the University of Arizona Cancer Center (Phoenix, AZ). All other cells were obtained from American Type Culture Collection (ATCC; Manassas, VA) and maintained in standard culture conditions at 37 °C in a humidified atmosphere with 5% CO₂. All cell culture media were obtained from Thermo Fisher Scientific, Waltham, MA. MIA PaCa-2 (CRM-CRL-1420) cells were cultured in Dulbecco’s Modified Eagle’s Medium (DMEM) high glucose supplemented with 10% fetal bovine serum (FBS) and 2.5% horse serum. PANC-1 (CRL-1469), UMUC3 and A549 cells were maintained in DMEM high glucose with 10% FBS. NCI-H520 (HTB-182), UACC903, UACC1113, 5637 (HBT9) and TCCSUP (HTB-5) cells were cultured in RPMI-1640 with 10% FBS. HEK293T (CRL-3216) cells, used for lentiviral production, were grown in high-glucose DMEM supplemented with 10% FBS, 1% glutamine, and 1% sodium pyruvate. All cell lines were routinely monitored for expected morphology and growth rates in addition to regularly being screened for mycoplasma contamination (Lonza MycoAlert Mycoplasma Detection Kit: LT07-318) which consistently tested negative. Authentication was performed using a short tandem repeat (STR) profiling panel (Identifiler kit, Life Technologies, Carlsbad, CA), and resulting profiles were compared against publicly available reference datasets from ATCC and DSMZ (https://www.dsmz.de/). All cell lines demonstrated correct STR identity based on database records.

### Fine-mapping and MPRA variant selection

To derive credible sets, we fine mapped previously published GWAS for PDAC^46^ (9,040 cases, 12,496 controls), melanoma^47^ (36,760 cases, 375,188 controls), lung^49,50^ (EUR; 29,266 cases, 56,450 controls, EAS; 21,658 cases, 150,676 controls) and bladder cancer^48^ (13,790 cases, 343,502 controls). We filtered summary statistics^46–50^ to include all available variants within ±500kb of the most significant 5p15.33 SNP for each cancer type. We then used MungeSummStats^94^ (v1.18.0) to lift genomic coordinates over to GRCh38 and standardized effect alleles to the minor allele (**Supplementary Table 1**). We then utilized the Sum of Single Effects (SuSiE) method^9^ (v0.14.2) susie_rss() function under default parameters to derive credible sets (CSs) with *L* = 10 (up to 10 possible causal variants per credible set). LD matrices for SuSiE were generated using PLINK^95^ (v2.0.0-a.7) from the 1000 Genomes Project Phase 3 (1kGP v3) dataset^51^, using genotypes from unrelated East Asian ancestry samples (EAS) for East Asian lung cancer summary statistics and unrelated European ancestry (EUR) samples for the remaining four GWAS. For our MPRA, we included CCVs from each of the identified SuSiE credible sets in addition to the following correlated variants (1kGP v3 EUR for PDAC, melanoma, bladder and lung EUR, EAS for lung EAS), using an LD threshold *r*^2^ ≥ 0.6 as a compromise between inclusivity and specificity:

1. LD *r*^2^ ≥ 0.6 relative to the SNP with the highest posterior inclusion probability (PIP) for an identified SuSiE credible set.
2. LD *r*^2^ ≥ 0.6 relative to any signal-tagging SNP identified by Chen and colleagues in 2021.^5^
3. LD *r*^2^ ≥ 0.6 relative to any signal-tagging SNP identified by Wang and colleagues in 2014.^4^

This approach resulted in 116 5p15.33 CCVs which were included in the MPRA panel (**Supplementary Figure 1**). In addition to these variants, we also included 44 additional variants from previous MPRAs and luciferase reporter assays as positive controls (**Supplementary Table 3**).

### Epigenetic annotation of CCVs

CCVs were annotated with functional scoring metrics from RegulomeDB^57^ (v2.2) and FORGEdb^58^ using the LDlink online tool^96^. CCVs were further annotated for predicted disruptions or creations of transcription factor binding motifs using motifbreakR^59^. motifbreakR utilized TF models from HOCOMOCO v11. We used the information content method and required motif matches to meet a maximum p-value of 10⁻⁴. Strong motifbreakR effects were defined as a difference between reference and alternate alleles scores greater than 0.7. We further annotated motifbreakR results with colocalization of matching TF ChIP-Seq data from the ReMap22^93^ database.

### MPRA library design

For each CCV, we designed oligonucleotides comprised of 145 bases of contextual sequence (hg38) centered on either the effect or non-effect GWAS alleles, in both forward and reverse orientations with a unique 12-mer barcode separated by contiguous KpnI and XbaI restriction sites (structured oligo sequence: 5’-ACTGGCCGCTTCACTG-145 bases-GGTACCTCTAGA-12 base tag-AGATCGGAAGAGCGTCG-3’) (Agilent Technologies, Santa Clara, CA). Barcode sequences were preselected from the publicly available FREE barcodes database^12^ for balanced GC content, minimal homopolymer runs, and reduced hairpin propensity. Additionally, we filtered barcodes to exclude any known mammalian miRNA seed sequences using miRbase^13^. For each variant, we designed six constructs (reference, alternate, and core 20 bp randomized in both forward and reverse orientations), with 35 unique barcodes per construct. Seven variants (rs11278847, rs11414507, rs3215401, rs34218850, rs28379291, rs3030832, rs57516180) failed sequence design due to the presence of KpnI and XbaI restriction sites within their contextual sequence.

### MPRA plasmid pool construction, transfection and sequencing

To clone MPRA oligonucleotide pools into the pMPRA2 delivery vector (Addgene, Watertown, MA, plasmid #49350), 10 fmol of reconstituted oligonucleotides were first amplified by emulsion PCR (ePCR) using Herculase II Fusion Polymerase (Agilent) with primers containing SfiI restriction sites. ePCR was performed with the Micellula DNA Emulsion & Purification Kit (EURx/CHIMERx, Milwaukee, WI). The amplified oligonucleotide pools were then ligated to Illumina adapter sequences using the KAPA HyperPrep Kit (KAPA Biosystems, Wilmington, MA) and quantified with the KAPA qPCR assay. Tag recovery rates were assessed via Illumina MiSeq v3 (150 bp, paired-end reads; Illumina, San Diego, CA). ePCR products were digested with SfiI (New England Biolabs, Ipswich, MA) and ligated into the complementary SfiI site of the pMPRA1 vector (Addgene plasmid #49349) using Electroligase (New England Biolabs, Ipswich, MA) at a 1:25 vector-to-insert molar ratio. The ligation product was digested with KpnI and XbaI (New England Biolabs, Ipswich, MA) between the 145 bp element and 12 bp tag sequence to enable ligation of a *luc2* open reading frame (ORF) cloned from the corresponding restriction sites of the pMPRA2 vector. The final ligation product was electrotransformed into ONESHOT electrocompetent cells (Thermo Fisher Scientific), ensuring that each construct was represented in at least ten independent transformations. The cloned library intended for transfection was validated as a single band on an agarose gel following KpnI digestion.

We performed MPRA transient transfections in eight cancer cell lines: PANC-1, MIA PaCa-2, UACC903, UACC1113, A549, H520, UMUC3, and 5637 (We used 5637 bladder cancer cells for the MPRA (rather than TCCSUP for CRISPRi) because 5637 cells are amenable to transient transfection). Each cell line was transfected in biological quadruplicate, with replicate cultures plated on separate days. For each transfection, the number of transfected cells exceeded the library complexity by more than 100-fold to ensure comprehensive representation of all sequences. 14 µg of MPRA plasmid was transfected to cell cultures using Lipofectamine 3000 (Thermo Fisher Scientific) according to the manufacturer’s protocol. Cell densities were optimized for each line to maximize transfection efficiency, which was verified by a parallel GFP transfection assay and fluorescence microscopy (no less than ∼ 50% transfection efficiency). Cells were harvested 48 hours post-transfection for RNA isolation.

Total RNA was extracted with the RNeasy Kit (QIAGEN, Hilden, Germany) and enriched for mRNA using the Poly(A)Purist MAG Kit (Thermo Fisher Scientific). Complementary DNA (cDNA) was synthesized using SuperScript III Reverse Transcriptase (Thermo Fisher Scientific). Unique 12-mer barcode sequences were amplified from cDNA with Q5 High-Fidelity DNA Polymerase (New England Biolabs) using primers containing Illumina TruSeq adapter sequences. Tag sequence libraries were prepared from both input DNA and cDNA using identical workflows. Sequencing was performed on a NovaSeq 6000 SP flow cell (Illumina, San Diego, CA) with 100 bp, dual-indexed, single-end reads at NCI’s Cancer Genomics Research Laboratory. For each cell line, sequencing depth ranged from 150 to 300 million reads per sample, providing sufficient coverage for downstream analyses (**Supplementary Table 17**).

### MPRA data analyses

Using FASTQ files generated from transfected plasmid DNA and RNA transcript sequencing, we quantified reads that perfectly matched each 12 bp tag sequence, along with downstream regions encompassing the XbaI recognition site and 20 bp of the 3′ *luc2* sequence. For each transfection, tag counts per million (TPM) were calculated by dividing the raw tag count by the total number of sequence-matching tag counts per million reads. To infer the transcriptional activity of each element, a single pseudocount was added to every TPM value and the RNA/DNA ratio for each tag was then computed by dividing its RNA TPM by the corresponding DNA TPM. For each transfection, the median ratio across all tags was calculated, and adjusted ratios were obtained by dividing each tag’s ratio by this median to ensure standardized scaling across replicates. Adjusted ratios were subsequently log₂-transformed to derive activity levels associated with each tested tag sequence.

Across all eight cell lines, at least 97.05% of barcode sequences were detected in both input DNA libraries and RNA samples. Reproducibility between transfections was assessed using Pearson correlation. After removal of a single outlier replicate per cell line, transcriptional activity levels showed high correlation across transfection replicates (median Pearson’s *r*; PANC-1 = 0.455, MIA PaCa-2 = 0.236, UACC903 = 0.852, UACC1113 = 0.903, A549 = 0.999, NCI-H520 = 0.956, UMUC3 = 0.981, 5637 = 0.964; n = 4 transfections per cell line;). We restricted analysis to barcodes with ≥100 reads in either RNA or DNA libraries, retaining 89.65% of designed tags. To evaluate the transcriptional impact of alternate (ALT) versus reference (REF) alleles, we applied a standard linear regression model adjusting for strand orientation (forward or reverse) as a binary covariate and biological replicate as a categorical covariate. Model significance was assessed using the robust sandwich-type variance estimate (Wald test) to account for potential heteroskedasticity in measurement error, as implemented in the R package sandwich. P-values for each linear model were corrected for multiple testing using the Benjamini–Hochberg procedure, applying a false discovery rate (FDR) threshold ≤ 0.01 to identify variants exhibiting significant transcriptional differences between alleles and scrambled controls.

To further assess whether tested elements displayed divergent activity relative to transcriptionally inert controls, the mean log₂(RNA/DNA TPM) ratio for each allele was compared to the distribution of log₂(RNA/DNA TPM) ratios from all scrambled control constructs within the same cell line. For each cell line, the interquartile range (IQR) of scrambled control activity was determined: alleles with mean log₂(RNA/DNA TPM) ratios above the third quartile (Q3) were classified as putative activators, whereas those below the first quartile (Q1) were classified as putative suppressors (**Supplementary Fig. 6**). CCVs exhibiting an FDR <0.01 allelic effect, an absolute allelic fold change ≥15% and classification as either activator or suppressor were defined as MPRA-functional for a given cell line.

### Tiled CRISPR interference (CRISPRi) proliferation screen design

sgRNAs targeting chr5:1,233,287–1,380,000 bp (hg19) were downloaded from the UCSC Genome Browser “CRISPR Targets” track. We also incorporated 992 inert and validated negative control sgRNAs in addition to sgRNAs targeting a panel of gene promoters selected based on susceptibility gene status for a given cancer type or noted as proliferation-essential across DepMap CRISPRi screening datasets^60,97^. CRISPRi oligonucleotides were synthesized using the SurePrint Oligonucleotide Synthesis Platform (Agilent) in the following configuration: 5′-AGGCACTTGCTCGTACGACG-CGTCTCACACCG-[sgRNA20nt]-GTTTCGAGACG-ATGTGGGCCCGGCACCTTAA-3′. A total of 27,340 sgRNAs were included in the final library, of which 25,778 tiled across the 5p15.33 region. For the 116 CCVs identified in fine-mapping, there was a median of 14 sgRNAs mapping to within ±50bp of each CCV, with one CCV (rs62651531) failing to have any within this window. rs2853672 had the most sgRNAs mapping within this window, with 39 (**Supplementary Table 5**).

### CRISPRi sgRNA pool generation

sgRNA oligonucleotides were PCR-amplified using flanking primer sites: Forward – 5′ AGGCACTTGCTCGTACGACG 3′; Reverse – 5′ ATGTGGGCCCGGCACCTTAA 3′. Amplified products were cloned into the pXPR_050 vector (Addgene #96925) using the same emulsion PCR strategy described above for MPRA. ePCR products were purified with a QIAquick PCR Purification Kit (QIAGEN). The pXPR_050 backbone was digested with Esp3I (NEB), gel-extracted to isolate the linearized vector, and ligated to the sgRNA ePCR amplicons using Electroligase (NEB). The ligation product was electroporated into ONESHOT electrocompetent cells in six independent reactions. After a one-hour recovery at 37 °C in S.O.C. medium (Invitrogen; 300 rpm), serial dilutions were plated to yield > 100× colony-forming units (CFUs) per sgRNA in the pool. Bulk cultures were incubated overnight (37 °C, agitation at 300 rpm, 16 h) to minimize selective expansion of individual clones. Plasmids were isolated via six parallel endo-free maxi preps (QIAGEN), pooled, quantified by Qubit High Sensitivity DNA Assay and NanoDrop 8000 (Invitrogen, Waltham, MA), and PCR-amplified. Resultant amplicons were then sequenced for QC (Illumina MiSeq v3, 100 cycles, single-end). FASTQ files were analyzed for sgRNA recovery and quality metrics. >99 % of input sgRNAs were recovered, confirming library complexity sufficient for lentiviral packaging.

### Lentiviral Packaging

HEK293T cells were plated and grown to ∼70 % confluency in six T175 flasks in virus production medium (high-glucose DMEM media supplemented with 10% FBS, 1% glutamine, and 1% sodium pyruvate). Transfection mixtures were prepared using lipofectamine 3000 (Invitrogen) to the manufacturer’s protocol, incubated 15 min at room temperature, and added dropwise to cells. After 6–8 h at 37°C, media was replaced with nutrient-rich DMEM. At 48 hours post-transfection (Day 3), viral supernatants were collected, centrifuged to remove debris, and filtered (0.45 µm). To concentrate virus, 2× PEG precipitation buffer (Millipore Sigma, Burlington, MA) was added and incubated overnight at 4 °C. On Day 4, viral particles were pelleted by centrifugation, resuspended in 1 mL aliquots of 1× PBS (Quality Biological, Gaithersburg, MD), and stored at −80 °C pending titration and transduction. Viral titer was quantified using Lenti-X GoStix Plus (Takara Bio, Kusatsu, Japan). 20 μL of viral sample were mixed with 80 µL of chase buffer, incubated 10 min at room temperature, and assessed using the GoStix Plus iOS app. The control (C) and test (T) band intensity ratio was mapped to batch-specific calibration curves via QR code, yielding an estimate of p24 concentration and infectious titer.

### CRISPRi screen transduction, cell culture and sequencing

CRISPRi screening experiments were performed across eight cell lines (MIA PaCa-2, PANC-1, UACC903, UACC1113, A549, H520, UMUC3, and TCCSUP) which had been transduced with lentivirus for dCas9-KRAB-ZIM3 and cultured from clonal subdivisions. CRISPRi experiments were transduced at a multiplicity of infection (MOI) of 0.3 to ensure single sgRNA integration per cell. Transductions were conducted at a scale sufficient to maintain ∼500× representation of each sgRNA throughout the screen.

Three biological replicates were performed with transductions performed over consecutive days. For each replicate, counted cells were resuspended with lentiviral supernatant and 8 µg/mL polybrene and plated at 1×10⁷ cells per 15-cm dish. After 24 h, cultures were washed and selected with 10 µg/mL puromycin until all cells in no-virus controls were eliminated (approximately 24–72 h). Post-selection, CRISPRi populations were expanded to generate sufficient material for both the initial time point (T_0_) and continued passaging until subsequent harvests at T_1_ (∼12.5 doublings) and T_2_ (25 doublings).

Genomic DNA (gDNA) was isolated using the Quick-DNA Genomic DNA Kit (Zymo, Irvine, CA). For each biological replicate, gDNA was harvested at the initial post-selection time point and again at approximately 12.5 and 25 population doublings. The full quantity of gDNA from each sample was used as input for the first round of sgRNA amplification (PCR1), distributing reactions such that each contained 1 µg gDNA to preserve library complexity. PCR1 products were purified using a 1:1 Ampure XP bead cleanup (Beckman Coulter, Brea, CA). A second amplification (PCR2) was performed using the same number of reactions as PCR1 to incorporate sample-specific barcodes and full Illumina adapter sequences. Each 20 µL PCR2 reaction contained 15 ng of PCR1 product and PCR2-specific primers and was run for five cycles under the same cycling parameters used for PCR1. Reactions were pooled and purified using the QIAquick PCR Purification Kit (QIAGEN). Library size (324 bp) was verified using a BioAnalyzer High Sensitivity DNA assay (Agilent). A 30 µL aliquot of each prepared library was submitted to the NCI Cancer Genomics Research Laboratory (Rockville, MD, USA) for sequencing on an Illumina NovaSeq SP flow cell.

### CRISPRi screen quality control and data analyses

Sequencing data from the tiled CRISPRi proliferation screen spanning 5p15.33 (eight cell lines; biological triplicates; T0 immediately after puromycin recovery, T1 at ∼12.5 cumulative doublings, T2 at ∼25 doublings) were processed from raw FASTQ files using the standard MAGeCK-VISPR workflow (v0.5.6) to quantify sgRNA counts for the input plasmid, baseline, and post-proliferation timepoints and to perform library and assay QC. Across plasmid controls, we observed high read mapping efficiency (72.28–73.25%), near-uniform sgRNA representation (median Gini index 0.052; range 0.0488–0.055), and minimal dropout (mean reads/sgRNA 859.48; mean missing sgRNAs 155.69; **Supplementary Table 6**), whereas cell-line libraries showed progressive selection-associated skew (Gini increasing from T0 to T2; **Supplementary Table 6**).For each sample, MAGeCK was used to compute sgRNA log2 fold changes (LFCs) and gene-level statistics with multiple-testing correction (FDR), including positive-control evaluation using 95 proliferation-essential genes (**Supplementary Tables 7 and 9**) and targeted aggregation of sgRNAs assigned to the *TERT* and *CLPTM1L* promoter regions (*TERT;* chr5:1,294,628–1,298,988: GH05J001293 and *CLPTM1L*: chr5:1,343,491– 1,346,621: GH05J001343; **Supplementary Table 5**). Downstream analysis was restricted to comparisons between individual timepoints and plasmid counts. Fine-mapped CCVs were linked to perturbations by intersecting sgRNAs within ±50 bp of each CCV (median 14 sgRNAs/CCV; **Supplementary Table 5**) and testing CCV-assigned sgRNA sets for depletion or enrichment (**Supplementary Tables 18&19**). Finally, proliferation-essential regulatory elements were inferred from sgRNA LFCs using CRISPR-SURF^62^(v1.0) to deconvolve localized peaks of depletion at FDR ≤ 0.05; datasets in which deconvolution failed to converge (specified in **Supplementary Results**) were excluded, and locus-level patterns were visually confirmed in IGV by concordant read coverage and dropout profiles across replicates and cell lines.

### Epigenetic silencing of SNP and VNTR loci by targeted CRISPRi

Individual sgRNAs corresponding targeting selected CCVs and both VNTRs were cloned into the pXPR_050 CRISPRi expression vector. For each sgRNA, complementary oligonucleotides containing the 20-bp spacer sequence and BsmBI-compatible overhangs (Forward: 5′-CACCG[N₍₂₀₎]-3′; Reverse: 5′-AAAC[N₍₂₀₎]C-3′) were synthesized (IDT) and annealed using a standard thermocycler-based annealing protocol. pXPR_050 was linearized by BsmBI (NEB) digestion and gel-purified prior to ligation. Annealed oligonucleotides were ligated into the digested backbone using T4 DNA ligase (NEB). Ligations were transformed into *E. coli* TOP10 chemically competent cells (Thermo Fisher Scientific) and plated on LB-ampicillin agar plates (Quality Biological). Single colonies were expanded overnight in selective LB medium, and plasmid DNA was isolated using the ZymoPURE Plasmid Miniprep kit (Zymo). To confirm correct sgRNA insertion, plasmid DNA from each clone was barcoded and sequenced using the Oxford Nanopore Rapid Sequencing and Barcoding Kit on a single Flongle flow cell (Oxford Nanopore, Oxford UK). Sequence-verified clones were expanded in 50mL cultures and purified using the EndoFree Maxiprep kit (QIAGEN) for downstream experiments.

Lentiviral particles encoding individual sgRNAs were generated by transient transfection of HEK293T cells. Briefly, 4×10⁶ HEK293T cells were plated in 10cm^2^ dishes in high-glucose DMEM supplemented with 10% FBS, pyruvate, and glutamine (Quality Biological). Cells were transfected the following day with the sgRNA expression construct, psPAX2 packaging plasmid (Addgene #12260), and MD2.G envelope plasmid (Addgene #12259) using Lipofectamine 3000 (Thermo Fisher Scientific) in accordance with the manufacturer’s recommendations. Media was replaced 24 h post-transfection. Viral supernatant was collected at 48 h, clarified by low-speed centrifugation, filtered (0.45 µm), and concentrated by overnight PEG precipitation at 4 °C. Viral pellets were resuspended in sterile PBS and stored at −80°C. For sgRNA delivery, 1×10⁵ PANC-1 or A549 cells stably expressing dCas9-KRAB-ZIM3 were seeded in 6-well plates in selective media containing blasticidin and polybrene. 25µl of concentrated virus was added to each well, and plates were centrifuged at 300 × g for 1 h to enhance transduction efficiency, followed by overnight incubation at 37 °C. Media was replaced the next day with blasticidin- and puromycin-containing growth medium, and cultured for 96 hours before harvesting total RNA.

Total RNA was extracted using a QIAcube connect instrument with the RNeasy mini kit (QIAGEN). 1 µg of total RNA was converted to cDNA with SuperScript III reverse transcriptase (Thermo Fisher Scientific). RT–qPCR was performed using a QuantStudio 7 Flex system. *TERT* and *CLPTM1L* expression relative to negative control sgRNAs was assessed using TaqMan assays (Thermo Fisher Scientific) (*TERT:* Hs00972656_m1 *CLPTM1L:* Hs00363947_m1) with Ct values normalized to *HPRT1* values from the same well (Hs99999909_m1): An unpaired t-test with Welch’s correction was used to test for statistical significance.

### Targeted PacBio Amplicon Sequencing of *CLPTM1L* Intron 9

Genomic DNA samples for HapMap trios (CEU panel for CEPH Utah residents with ancestry from Northern and Western Europe, n = 72) and 1000 Genomes Project Europeans (Toscani in Italia (TSI); n = 114, Finnish in Finland (FIN); n = 100, British from England and Scotland (GBR); n = 100; Iberian Populations in Spain (IBS); n =100) were purchased from the Coriell Institute of Medical Research Institute (Camden, NJ). Genomic DNA samples for pancreatic cancer cases and controls were sourced from the first two phases of the PanScan GWAS ^20,21^.

Amplicons spanning chr5:1,325,380–1,330,639 (GRCh38) were generated using barcoded M13 primers and the PacBio SMRTbell prep kit v3.0 with minor protocol modifications. PCR cycling consisted of 30 cycles of 98 °C denaturation (10 s) and combined annealing/extension (6 min), followed by a 10 min final extension at 72 °C. Amplicons were purified with AMPure beads, quantified by PicoGreen (Invitrogen), barcoded with an additional 10 PCR cycles, pooled, and sequenced on a Sequel II SMRT Cell. HiFi reads were processed using SMRT Analysis for circular consensus sequence (CCS) generation and demultiplexed using Lima.

### Genotyping of VNTRs and SNPs from HiFi reads

HiFi amplicon reads were aligned to GRCh38 using pbmm2 with a minimum read-length threshold of 4,700 bp. Aligned HiFI reads were then processed with PacBio Amplicon Analysis^98^ (pbAA). Reads were first clustered to the target region (amplicon ±100 bp; chr5:1325280–1330739) using pbAA cluster, and haplotype tags (HP) were assigned to each read using pbAA bampaint. For each sample, the two modal clusters (corresponding to diploid haplotype sequences) were retained, and reads from minor artefactual clusters were excluded. Consensus sequences for each haplotype were reconstructed, realigned to GRCh38, and used as input for tandem repeat calling with TRGT^99^ and SNP genotyping with DeepVariant/GLnexus^100^ followed by read-based phasing of VNTRs and SNPs using HiPhase^101^.

We assessed genotyping concordance across 484 1kGP EUR samples between PacBio-derived SNP calls and orthogonal short read sequence calls^51^. For 17 SNPs passing quality thresholds (MAF_1kGP_ ≥ 0.01; HWE P_1kGP_ ≥ 0.05), minor allele frequencies (MAF) derived from PacBio SNP calls were highly concordant with those from 1kGP short-read WGS calls (Pearson’s *r* = 0.99; **Supplementary Table 10**), with VNTR genotypes also conforming to Mendelian segregation in 24 HapMap CEU trios (**Supplementary Table 11**).

For TRGT VNTR genotypes, we derived four complementary allele-encoding approaches:

1. Discrete VNTR alleles were maintained as distinct sequences denoted by their length and assigned a unique index number (<LEN_N>).
2. Distinct VNTR alleles described above were grouped together based on length irrespective of their sequence composition (<LEN>)
3. VNTR lengths and their associated allele counts were used to assign lengths based on k-means clustering (4 clusters for VNTR1, 3 for VNTR2), (<Cluster_n>).
4. Binary long/short VNTR genotypes were derived by combining (<L/S>).

CEU trios and EUR samples were analyzed jointly. Of 503 sequenced samples, 481 were matched to individuals in the 1000 Genomes Phase 3 high-coverage WGS dataset. Mendelian transmission was evaluated across 24 CEU trios using both VNTR1 and VNTR2 consensus alleles with all trios demonstrating complete Mendelian concordance. To further validate genotyping accuracy, variant calls for 31 dbSNP 155-annotated variants were compared with orthogonal 1000 Genomes short-read genotypes^51^. PacBio SNP genotypes demonstrated negligible differences in MAF (Median ΔMAF= 0.015, range = 0.003-0.228) between PacBio and 1000G WGS genotypes **(Supplementary Table 11),** indicating that accompanying VNTR calls were reflective of true genotypes.

### VNTR imputation and association analysis

To impute VNTR alleles into the broader PanScan GWAS^46^, we generated a reference panel designed to only impute VNTR alleles whilst leveraging phase information from both PacBio-genotyped SNPs and accurately-imputed variants from the same samples. Using data from the PanScan 940 PDAC cases and 870 controls subjected to amplicon sequencing, we compiled a multisample VCF file comprised of all VNTR genotypes (each genotyping method represented as a distinct multiallelic row to be imputed as a separate variant) along with corresponding SNP calls that met MAF > 1%, call rate = 1, and Hardy–Weinberg equilibrium P > 10⁻⁵ (11 SNPs total). Next, we took imputed dosages from the same sample set which had been imputed using the TOPMed imputation server, excluding any variants outside of chr5:0-6Mb, MAF < 1%, imputation R2 < 0.9 and any overlap with SNPs already genotyped by PacBio. VCFs were concatenated into a single file, sorted and then phased using SHAPEIT4.2^102^, leveraging HiPhase-derived phase set tags (PS) as a haplotype scaffold to preserve physical phasing information. Recipient PanScan genotypes were prepared similarly to the 1,810-reference panel cohort, filtering for position, MAF and imputation R2. To accommodate downstream imputation and association frameworks, multiallelic VNTR sites were decomposed to biallelic variants using bcftools^103^. VNTR alleles were then imputed into three cohorts of recipient samples (PanScan I&II (additional 2,585 cases, 2,823 controls)^66,67^, PanScan III (1,597 cases, 4,799 controls)^52^, and PanC4 (from the Pancreatic Cancer Case-Control Consortium^46^, dbGaP Study Accession: phs000648.v2.p1; 3,884 cases, 3586 controls; approval to Laufey T. Amundadottir)) using Beagle^104^ (v5.4) whilst retaining hard called VNTR alleles in the 1,810 reference samples for downstream regression. Downstream analysis of linkage disequilibrium and logistic regression and conditional analyses were performed using PLINK^95^ (v2.0.0-a.7). For logistic regression, we analyzed each cohort independently, including age, sex, sample collection site and eigenvectors from principal components analysis as fixed covariates as performed in the original meta-analysis^46^. We then meta-analyzed each cohort using PLINK (v1.9).

### Analysis of VNTR-transcription factor binding

To assess VNTR-enriched transcription factor binding sites, we obtained VNTR1 sequence in FASTA format from the UCSC genome browser simple repeats track (GRCh38). VNTR1 was then analyzed with the TRAP^69^ (Transcription Factor Affinity Prediction) online tool. TRAP provides an enrichment measure indicative of whether a transcription factor binds the VNTR sequence more strongly than expected relative to human promoters curated by TRANSFAC (**Supplementary Table 15**). We subsequently visualized VNTR1 RU composition and TEAD motif enrichment using motifscope^105^.

To validate *in*-*silico* TF binding for VNTR1, we designed biotinylated double-stranded oligonucleotides corresponding to the VNTR1 reference repeat unit (RU) and a +15-bp shifted RU were synthesized (IDT; **Supplementary Table 20**). Protein–DNA pulldown and mass spectrometry analyses were performed at the National Cancer Institute’s Protein Characterization Laboratory (Frederick, MD). Oligonucleotides were annealed in 2× binding and wash buffer (10 mM Tris pH 7.5, 1 mM EDTA, 2 M NaCl) and incubated with Neutravidin beads for immobilization. For each pulldown, 200 pmol of duplex oligo was incubated with 400 µg of nuclear extract prepared from MIA PaCa-2 cells in IP buffer (50 mM Tris pH 8.0, 150 mM NaCl, 0.1% NP-40, 1 mM MgCl₂, 100 µM ZnSO₄, 1 mM DTT, 5% glycerol, protease inhibitors). Competition controls included 50× and 100× excess of unlabeled duplex DNA to assess binding specificity. Following a 1hour incubation with oligo-conjugated beads, samples were washed in IP buffer, resuspended in SDS loading buffer, and submitted for mass spectrometry. Peptide identification and quantification followed established core facility pipelines. Relative peptide abundance across competitor and non-competitor conditions was compared using a one-sided *t*-test to determine significant displacement of DNA-associated proteins.

### VNTR repeat unit (RU) protein binding analysis by DNA pulldown

Biotinylated double-stranded DNA probes were generated by annealing complementary single-stranded oligonucleotides (100 μM each strand, 3 μl per strand, 300 pmol total, IDT, **Supplementary Table 20**) comprising sequences representative of the 1 reference RU and 15bp shifted sequence as used for global proteomics. Annealed probes were immobilized on NeutrAvidin agarose beads (30 μl) and incubated with nuclear protein extracts (100 μg) prepared using the NE-PER Nuclear and Cytoplasmic Extraction Kit (Thermo Fisher Scientific, Waltham, MA). Following incubation, protein-DNA-bead complexes were pelleted by centrifugation, and both the supernatant and bead-bound fractions were collected for analysis.

Proteins were eluted from the beads by heating in sample buffer. Input nuclear extract (20 μg), supernatant fractions (20 μg), and bead-bound proteins were resolved on Criterion XT 3-8% Tris-Acetate precast gels (Bio-Rad #3450129, Hercules, CA) using XT Tricine SDS running buffer (Bio-Rad #1610790) optimized for separation of 40-400 kDa proteins. Proteins were transferred to 0.45 μm pore PVDF membranes (Thermo Fisher Scientific #88518) using standard wet transfer methods. Membranes were stained with 0.1% Ponceau S (G Biosciences #786-575, VWR #89167-800) to verify transfer efficiency.

Membranes were blocked in 2% bovine serum albumin (BSA) in TBST (10 mM Tris-HCl pH 7.5, 150 mM NaCl, 0.05% Tween-20) and incubated overnight at 4°C with primary antibodies diluted 1:1,000 in 2% BSA/TBST: rabbit polyclonal anti-SP1 (Abcam ab13370), rabbit polyclonal anti-TAZ (Abcam ab110239), rabbit monoclonal anti-TEAD1 (Abcam ab133533), mouse monoclonal anti-TEAD1 (BD Transduction 610923), rabbit polyclonal anti-TEAD4 (Thermo Fisher Scientific 12418-1-AP), mouse monoclonal anti-TEAD4 (Santa Cruz sc-101184), rabbit monoclonal anti-YAP1 (Abcam ab52771), and rabbit monoclonal anti-YY1 (Abcam ab109228). Following TBST washes, membranes were incubated for 1 h at room temperature with HRP-conjugated secondary antibodies diluted 1:2,000 in 2% BSA/TBST: donkey anti-rabbit HRP (Abcam ab205722) or donkey anti-mouse HRP (Abcam ab7061). Chemiluminescence was detected using SuperSignal West Femto Maximum Sensitivity Substrate (Thermo Fisher Scientific) and imaged on a ChemiDoc imaging system (Bio-Rad).

### VNTR luciferase assays

To assess VNTR1 transcriptional activity, VNTR1 alleles from samples from the 1000 Genomes European Ancestry Set were amplified using custom PCR primers (**Supplementary Table 20)**, allowing for Gateway cloning. HapMap samples HG00171 (2,586 bp), GM20759 (2,789 bp), GM20818 (2,933 bp) and GM20787 (3,566 bp) were selected to represent both the extremes of allelic length distribution and the two modal lengths. A *negative* control construct was generated by amplifying a 2.7 kb region in intron 9 of *TERT* which was not annotated by any epigenetic marks in ENCODE or any CRISPR-SURF peaks from the screen. PCR, Gateway cloning and sequence verification were performed at the National Cancer Institute’s Protein Expression Laboratory (Frederick, MD, USA) using a protocol described by Wall et al.^26^

PANC-1 and A549 cell lines were seeded in 48-well plates at densities of 2 × 10⁴ and 4 × 10⁴ cells per well, respectively. Cells were transfected the following day with firefly luciferase reporter constructs using Lipofectamine 3000 (Invitrogen), together with 250 ng of the corresponding control plasmid and 2.5 ng of a Renilla luciferase plasmid (pGL4.74). Each construct was assayed in biological triplicate with six technical replicates per experiment. Transfection media was replaced after 6 h with complete growth medium, and cells were lysed 24 h later in passive lysis buffer (Promega Madison, WI). Firefly and Renilla luciferase activities were quantified using the Dual-Luciferase Reporter Assay System on a GloMax Explorer plate reader (Promega) following the manufacturer’s protocol. For Hippo pathway inhibition studies, reporter assays were performed in the presence of 0.5, or 1.0 μM verteporfin (Millipore-Sigma), alongside a DMSO vehicle control. Firefly luciferase values were normalized to Renilla luciferase for each well, averaged across technical replicates, and expressed relative to the activity of the negative control plasmid. Data were pooled across experiments, and mean values with standard deviations were plotted using ggplot2. Statistical comparisons between constructs comprised a pairwise two-sided unpaired Welch’s t-test.

### Reagents and oligonucleotides

All reagents, oligonucleotides, plasmids, antibodies, commercial kits, and DNA samples used in this study, including supplier information and catalog numbers, are listed in **Supplementary Table 20**.

## Supplementary Results

### MPRA functional score enrichment

Several databases based on publicly available epigenomics data exist for the functional annotation of GWAS SNPs purporting to aide in triaging of functional variants from correlated proxies. To investigate the predictive power of these tools, we assessed whether annotation scores were enriched for MPRA-validated functional variants. For each set of MPRA variants (CCVs and positive controls), we assayed SNPs stratified by cell line and in a combined model and performed Fisher’s exact tests to determine whether specific RegulomeDB and FORGEdb scores were associated with MPRA hit status. We evaluated multiple definitions of functional significance: (i) statistical significance alone (FDR ≤ 0.01), (ii) statistical significance with meaningful effect size (FDR ≤ 0.01 and absolute fold change ≥ 15%), (iii) active regulatory element status (activator or suppressor designation in either allele), and (iv) the intersection of all criteria. Annotative scores were found to have limited predictive capacity in for MPRA status, with the only significant associations observed between active element status and FORGEdb scores of 8 in A549 (*P*_adj_ = 0.0257) regulomeDB scores of 1b in PANC-1 cells (*P_adj_* = 0.0469). These findings suggest that annotative scores should be used on a cautiously across individual loci and be used as a complement to comprehensive functional screening.

### CRISPRi Screening Validation

Sequencing data from the tiled CRISPRi proliferation screen of 5p15.33 across eight cell lines were processed using MAGeCK-VISPR^106^ (v0.5.6) for sgRNA-level and positive control QC. FASTQ files corresponding to the plasmid library, baseline (T₀), and post-proliferation (T₁ and T₂) timepoints were analyzed using the standard MAGeCK-VISPR workflow to assess library quality, read mapping, and assay efficacy. Across all plasmid control libraries, high mapping efficiency (72.28%–73.25%) and uniform sgRNA representation were observed, with Gini indices ranging from 0.05 to 0.06, consistent with even distribution of sgRNA counts. In contrast, cell-line derived libraries exhibited selection-mediated variability through timepoints, reflected by progressive elevation of Gini indices—from 0.05–0.10 at T₀ to 0.13 at T₁ and up to 0.17 at T₂.

To evaluate assay sensitivity and reproducibility, depletion and enrichment statistics were derived from the normalized sgRNA counts. Enrichment of small P-values was noted across positive-control proliferation genes (**Supplementary Table 7**), including *KRAS*, *MYC*, and *CCND1*, indicative of proliferation-mediated of associated sgRNAs. Consistent and significant depletion was observed for sgRNAs targeting the *TERT* and *CLPTM1L* promoter regions, defined as chr5:1,294,628–1,298,988 (GeneHancer ID: GH05J001293) and chr5:1,343,491–1,346,621 (GeneHancer ID: GH05J001343), respectively, encompassing 915 and 632 intersecting sgRNAs. These loci exhibited strong depletion signals across multiple cell lines, consistent with their established roles in supporting proliferative capacity (**Supplementary Table 5**).

Inference of proliferation-essential regulatory elements from the tiling data was performed using CRISPR-SURF^62^ (v1.0) which applies a deconvolution-based approach to aggregate sgRNA-level effects into distinct regions of proliferation-essentiality. CRISPR SURF deconvolution successfully resolved high-confidence peaks of depletion overlapping the *TERT* and *CLPTM1L* promoters in six of eight cell lines. Convolution failed to converge for MP2 (T₁), UACC903 (all timepoints), and H520 (T₁–T₂) datasets, which were excluded from downstream integration analyses. Visual inspection of aligned read coverage and sgRNA depletion profiles in Integrative Genomics Viewer (IGV) confirmed reproducible patterns of promoter-specific dropout across cell lines. Together, these analyses demonstrated that the CRISPRi proliferation screens achieved robust coverage, reproducible sgRNA representation, and consistent detection of proliferation-essential loci, including canonical oncogenes and core promoter elements within the 5p15.33 region.

For the 116 CCVs identified in fine mapping, there was a median of 14 sgRNAs intersecting within ± 50bp of each CCV, with one CCV (rs62651531) failing to have any within this window. rs2853672 had the most intersecting sgRNAs, with 39 (**Supplementary Table 5**). We annotated 915 and 632 of the tiling sgRNAs as intersecting the *TERT* and *CLPMT1L* promoters, respectively (Methods). We transduced lentiviral packaged sgRNAs into the same cell lines used for MPRA which were clonally engineered to stably express dCas9-KRAB-ZIM3 (**Supplementary Table 4**). Screens were performed in biological triplicate for 25 cumulative cell doublings and harvested at an initial time point (T_0_, immediately following puromycin selection and initial recovery), T_1_ (∼12.5 cumulative screen doublings), and T_2_ (25 screen doublings).

Following sequencing of sgRNAs from harvested time points and the primary plasmid, we assessed library quality, read mapping, and assay efficacy using the output of MAGeCK. Sequencing libraries from input plasmid and cell line demonstrated high mapping efficiency, even sgRNA read distribution and minimal sgRNA dropout (Average reads/sgRNA = 859.48, average missing sgRNAs = 155.69, **Supplementary Table 6**). Gini indices, a measure of sgRNA read distribution across a sample, increased progressively through plasmid (median Gini = 0.052, range = 0.0488-0.055) and cell line timepoints from (range T₀ = 0.065, 0.0524-0.175, T₁ = 0.079, 0.055-0.1548, and T₂ = 0.085, 0.052-.1743), indicative of CRISPRi-mediated negative selection of sgRNAs over time.

We used MAGeCK to calculate log_2_ fold changes for each timepoint relative to the plasmid in addition to its preceding timepoint and measured inter-replicate Pearson correlation to assess reproducibility (**Supplementary Fig. 6**). Across all comparisons (three replicate pairs per experiment), the inter-replicate Pearson correlation coefficients ranged from –0.42 to 1.00, with a median of 0.46 (IQR 0.20–0.81), indicating that while some comparisons show very strong agreement between replicates, others are considerably noisier. Reproducibility was highest for the plasmid control samples. For plasmid experiments the mean inter-replicate correlation was 0.78 (median 0.86), and ∼76% of replicate pairs had *r* > 0.7. In contrast, cell line derived time-point contrasts were less consistent: T_1__vs_T_0_ / T_2__vs_T_0_ comparisons across all cell lines had a median *r* of 0.20. T_2__vs_T_1_ comparisons were lower still (median 0.05), with several replicate pairs near zero or weakly negative. There was also a clear trend with time: at T0, replicate correlations were generally high (mean 0.68, median 0.82), decreasing at T_1_, (median *r* = 0.54) and further at T_2_, with a median *r* =0.27. By cell line, PANC1, TCC, UMUC3, A549 and MP2 showed the best overall replicate concordance (median *r* range across timepoints = 0.54–0.80), whereas UACC903, H520 and UACC1113 were more variable (median r = 0.15–0.29). With this in mind, we restricted downstream analysis only to comparisons between sgRNA counts derived from cell line time points and their corresponding count in the input plasmid.

To assess the efficacy of CRISPRi systems, we assessed knockdown of 95 genes selected for proliferation-essentiality and cancer-type susceptibility status using MAGeCK. Across all timepoints and cell lines, we observed a median of 39 knocked down genes at FDR ≤ 0.05 (41.05%, range 5-60, **Supplementary Tables 7 and 9**). Depleted genes varied considerably between cell lines and cancer types, suggestive of divergent physiology. Ranking each gene within a timepoint in ascending order of depletion FDR, we found top ranked associations for canonical oncogenes such as *KRAS* and *CCND1* in most cell lines across time points (**Supplementary Table 7**).

Depletion testing of both genes in UACC903 melanoma cell lines was consistent with subsequent confirmation of progressive reduction in dCas9-KRAB-ZIM3 expression (**Supplementary Fig. 7**). We noted no significant knockdown of the canonical tumor suppressor *TP53*, concordant with the depletion-based design of our screen.

To further validate our experimental design, we assigned sgRNAs from the tiling component of the pool to either the *TERT* or *CLPTM1L* promoter and tested both genes using MAGeCK Analysis of *TERT* promoter targeting-sgRNAs demonstrated striking cell line-specific effects, suggestive of both physiological differences and experimental limitations. NCI-H520 lung cancer cells showed the most dramatic depletion of *TERT* sgRNAs, with log_2_ fold changes progressing from −0.028 at T_0_ to −1.297 at T_2_ (FDR = 3.30×10⁻⁵), indicating strong growth dependency on the *TERT* promoter. A549 cells exhibited more modest but consistent depletion of *TERT* promoter sgRNAs across timepoints (log_2_ fold change = −0.062, FDR = 2.55×10⁻⁴ at T_2_). Among melanoma lines, UACC1113 demonstrated significant growth dependency on the *TERT* promoter (log_2_ fold change= −0.217, FDR = 1.90×10⁻⁵ at T_2_), while UACC903 showed minimal effects across all timepoints (maximum 7 total depleted positive control genes). Bladder cancer line 5637 transitioned from modest enrichment at T_0_ (LFC = 0.030) to significant depletion by T_2_ (LFC = −0.082, FDR = 3.30×10⁻⁵), whereas UMUC3 maintained consistent low-level enrichment (LFC ∼0.04) with marginal significance. Pancreatic lines showed variable patterns: MIA PaCa-2 exhibited minimal fold changes but significant FDRs, while PANC-1 progressed from non-significant enrichment at T0 to moderate depletion at later timepoints (LFC = −0.080, FDR = 2.10×10⁻⁵ at T2). *CLPTM1L* promoter silencing generally paralleled *TERT* promoter effects but with attenuated magnitude. NCI-H520 again displayed the strongest dependency (LFC = −0.703, FDR = 4.32×10⁻⁴ at T_2_), followed by UACC1113 (LFC = −0.167, FDR = 1.14×10⁻³ at T_2_). Notably, several samples showed significant *CLPTM1L* promoter effects at early timepoints: UACC1113 at T_0_ (FDR = 4.32×10⁻⁴) and MIA PaCa-2 at T_0_ (FDR = 4.50×10⁻⁵), suggesting immediate growth consequences upon *CLPTM1L* promoter perturbation in these contexts. A549 showed modest depletion (LFC = −0.059 at T2), while UMUC3 and PANC-1 displayed weak or non-significant effects (**Supplementary Table 8**).

We then went on to apply MAGeCK to 5p15.33 CCVs, assigning sgRNAs to CCVs by intersecting them with ± 50 bp each fine mapped CCV. SNP-level analysis identified 5 to 54 significantly depleted variants per sample, with depletion counts generally tracking overall screen sensitivity (**Supplementary Table 9).** NCI-H520 yielded the highest SNP hits at T_2_ (n=54), coinciding with maximal promoter effects and suggesting coordinated regulatory dependencies across the locus. 5637 and A549 maintained consistent SNP depletion across time points while melanoma line UACC903 showed minimal regulatory variant sensitivity (n=5 across all timepoints), consistent with its weak overall selection. PANC-1 demonstrated a notable increase in depleted SNPs from T_0_ (n=10) to T_1_/T_2_ (n=35), suggesting that SNP-mediated *cis-*regulatory effects may take a longer period to manifest in phenotypic divergence. While our we primarily hypothesized that functional SNPs would occupy loci which would be epigenetically silenced upon CRISPRi inhibition, we observed six SNPs for which sgRNAs were significantly enriched (**Supplementary Table 10).**

Next, we used CRISPR-SURF^8^ to deconvolve sgRNA log_2_ fold changes for each timepoint into discrete regions of putative functional regulatory activity. Unlike gene-centric or SNP-focused analyses, CRISPR-SURF operates in an unbiased, genome-agnostic manner, identifying functional regulatory regions based solely on local enrichment of depleted sgRNAs without prior assumptions about annotated features or genetic markers. This approach enabled discovery of both promoter-proximal and distal regulatory elements, as well as unannotated functional sequences across the tiled 5p15.33 region (**Supplementary Fig. 9**). CRISPR-SURF deconvolution failed for MIA PaCa-2 T_1_ and NCI-H520 T_1_, and T_2_.and UACC903 T_0._ Of the remaining datapoints, we identified a total of 2,718 proliferation-essential elements at FDR ≤ 0.05, with a median length of 470 bp.

## Acknowledgments

This research was supported in part by the Intramural Research Program of the National Institutes of Health (NIH). The contributions of the NIH authors are considered Works of the United States Government. The findings and conclusions presented in this paper are those of the authors and do not necessarily reflect the views of the NIH or the U.S. Department of Health and Human Services

The authors acknowledge the research contributions of the NCI Cancer Genomics Research Laboratory for their expertise, execution, and support of this research in the areas of project planning, wet laboratory processing of specimens, and bioinformatics analysis of generated data. We thank Dr. Raj Chari, Director, Genome Modification Core, Frederick National Laboratory for Cancer Research, National Cancer Institute, Frederick, MD, for assistance with CRISPR guide design. We also thank Dr. Dominic Esposito, Director, Protein Expression Laboratory (PEL) Frederick National Laboratory for Cancer Research, National Cancer Institute, Frederick, MD, for assistance in cloning of VNTR luciferase constructs.

This work utilized the computational resources of the NIH HPC Biowulf cluster (https://hpc.nih.gov).

SL was supported by CA297773 (NIH).

## The Pancreatic Cancer Cohort Consortium and the Pancreatic Cancer Case-Control Consortium (PanScan/PanC4) Acknowledgements and Funding statements

**American Cancer Society**. The authors express sincere appreciation to all Cancer Prevention Study-II participants, and to each member of the study and biospecimen management group. The authors would like to acknowledge the contribution to this study from central cancer registries supported through the Centers for Disease Control and Prevention’s National Program of Cancer Registries and cancer registries supported by the National Cancer Institute’s Surveillance Epidemiology and End Results Program. The American Cancer Society funds the creation, maintenance, and updating of the Cancer Prevention Study-II cohort.

The **ATBC Study** was supported by the Intramural Research Program, Division of Cancer Epidemiology and Genetics of the U.S. National Cancer Institute (NCI), National Institutes of Health.

Cancer incidence data for **CLUE** were provided by the Maryland Cancer Registry, Center for Cancer Surveillance and Control, Department of Health and Mental Hygiene, 201 W. Preston Street, Room 400, Baltimore, MD 21201, http://phpa.dhmh.maryland.gov/cancer, 410-767-4055. We acknowledge the State of Maryland, the Maryland Cigarette Restitution Fund, and the National Program of Cancer Registries of the Centers for Disease Control and Prevention for the funds that support the collection and availability of the cancer registry data.” We thank all the CLUE participants.

## EPIC-Europe Funding Statement and Disclaimer

EPIC Financial support: The coordination of EPIC-Europe is financially supported by International Agency for Research on Cancer (IARC) and also by the Department of Epidemiology and Biostatistics, School of Public Health, Imperial College London which has additional infrastructure support provided by the NIHR Imperial Biomedical Research Centre (BRC).

The national cohorts are supported by: Danish Cancer Society (Denmark);Ligue Nationale Contre le Cancer, Institut Gustave Roussy, Mutuelle Générale de l’Education Nationale (MGEN), Institut National de la Santé et de la Recherche Médicale (INSERM), French National Research Agency (ANR, reference ANR-10-COHO-0006), French Ministry for Higher Education (subsidy 2102918823, 2103236497, and 2103586016) (France); German Cancer Aid, German Cancer Research Center (DKFZ), German Institute of Human Nutrition Potsdam-Rehbruecke (DIfE), Federal Ministry of Research, Technology and Space (BMFTR) (Germany); Associazione Italiana per la Ricerca sul Cancro-AIRC-Italy, Italian Ministry of Health, Italian Ministry of University and Research (MUR), Compagnia di San Paolo (Italy); Dutch Ministry of Public Health, Welfare and Sports (VWS), the Netherlands Organisation for Health Research and Development (ZonMW), World Cancer Research Fund (WCRF), (The Netherlands); UiT The Arctic University of Norway; Health Research Fund (FIS) - Instituto de Salud Carlos III (ISCIII), Regional Governments of Andalucía, Asturias, Basque Country, Murcia and Navarra, and the Catalan Institute of Oncology - ICO (Spain); Swedish Cancer Society, Swedish Research Council of Region Skåne and Region Västerbotten (Sweden); Cancer Research UK (C864/A14136 to EPIC-Norfolk; C8221/A29017 to EPIC-Oxford), Medical Research Council (MR/N003284/1, MC-UU_12015/1 and MC_UU_00006/1 to EPIC-Norfolk (DOI 10.22025/2019.10.105.00004); MR/Y013662/1 to EPIC-Oxford) (United Kingdom). Previous support has come from “Europe against Cancer” Programme of the European Commission (DG SANCO).

EPIC Disclaimer: Where authors are identified as personnel of the International Agency for Research on Cancer / World Health Organization, the authors alone are responsible for the views expressed in this article and they do not necessarily represent the decisions, policy or views of the International Agency for Research on Cancer / World Health Organization.

**The Health Professionals Follow-up Study** is supported by NIH grant UM1 CA167552. from the National Cancer Institute, Bethesda, MD USA.

The work at **Johns Hopkins University** was supported by the NCI Grants P50CA062924 and R01CA154823. Additional support was provided by the Lustgarten Foundation, Susan Wojcicki and Dennis Troper and the Sol Goldman Pancreas Cancer Research Center.

The **Multiethnic Cohort Study (MEC)** is supported by NCI grant U01CA164973.

The **MD Anderson case control study** was supported by NIH R01 CA98380.

The **Melbourne Collaborative Cohort Study (MCCS)** cohort recruitment was funded by VicHealth and Cancer Council Victoria. The MCCS was further augmented by Australian National Health and Medical Research Council grants 209057, 396414 and 1074383 and by infrastructure provided by Cancer Council Victoria. Cases and their vital status were ascertained through the Victorian Cancer Registry and the Australian Institute of Health and Welfare, including the National Death Index and the Australian Cancer Database.

Work at **Memorial Sloan Kettering Cancer Center** was funded by the center grant P30CA008748 (Selwybn Vickers).

The **NYU-WHI** study was funded by NIH R01 CA098661, UM1 CA182934 and center grants P30 CA016087 and P30 ES000260.

The **Nurses’ Health Study** is supported by NIH grants UM1 CA186107, P01 CA87969, and R01 CA49449 from the National Cancer Institute, Bethesda, MD USA.

The **Physicians’ Health Study** was supported by research grants CA-097193, CA-34944, CA-40360, HL-26490, and HL-34595 from the National Institutes of Health, Bethesda, MD USA

The **Queensland Pancreatic Cancer Study** was supported by a grant from the National Health and Medical Research Council of Australia (NHMRC) (Grant number 442302). RE Neale is supported by a NHMRC Senior Research Fellowship (#1060183).

The **SELECT** study is supported by National Institutes of Health grant award number UG1CA189974 (CD Blanke), and U01 CA182883 (CM Tangen/IM Thompson). The authors thank the site investigators and staff and, most importantly, the participants who donated their time to this trial.

Data collection at the **University of Toronto** was supported by the CIHR Canada Research Chair and from the Canadian Cancer Society Research Institute.

The **WHI** program is funded by the National Heart, Lung, and Blood Institute, National Institutes of Health, U.S. Department of Health and Human Services through contracts 75N92021D00001, 75N92021D00002, 75N92021D00003, 75N92021D00004, 75N92021D00005.The authors thank the WHI investigators and staff for their dedication, and the study participants for making the program possible. A full listing of WHI investigators can be found at: https://www-whi-org.s3.us-west-2.amazonaws.com/wp-content/uploads/WHI-Investigator-Short-List.pdf.

The **Harvard Women’s Health Study** funding was as follows: CA047988, CA182913, HL043851, HL080467 and HL099355.

The **Yale (CT) pancreas cancer study** is supported by National Cancer Institute at the U.S. National Institutes of Health, grant 5R01CA098870. The cooperation of 30 Connecticut hospitals, including Stamford Hospital, in allowing patient access, is gratefully acknowledged. The Connecticut Pancreas Cancer Study was approved by the State of Connecticut Department of Public Health Human Investigation Committee. Certain data used in that study were obtained from the Connecticut Tumor Registry in the Connecticut Department of Public Health. The authors assume full responsibility for analyses and interpretation of these data.

## Consortia authors from the Pancreatic Cancer Cohort Consortium and Pancreatic Cancer Case-Control Consortium (PanScan/PanC4)

Jun Zhong^1^, Demetrius Albanes^2^, Gabriella Andreotti^3^, Samuel O. Antwi^4^, Alan A Arslan^5^, William R. Bamlet^6^, Laura Beane-Freeman^3^, Sonja I Berndt^3^, Paige M. Bracci^7^, Daniele Campa^8^, Federico Canzian^9^, Stephen J Chanock^10^, Yu Chen^11^, Charles C Chung^12^, Mengmeng Du^13^, A. Heather Eliassen^14,15,16^, Steven Gallinger^17^, J. Michael Gaziano^18,19^, Jeanine Genkinger^20^, Michael Goggins^21^, Phyllis J Goodman^22^, Christopher A Haiman^23^, Manal Hassan^24^, Belynda Hicks^12^, Rayjean J. Hung^25^, Amy Hutchinson^12^, Verena Katzke^26^, Manolis Kogevinas^27,28^, Charles Kooperberg^29^, Martijn P Kolijn^30,31^, Peter Kraft^32^, Robert C. Kurtz^33^, I-Min Lee^34,14^, Loic LeMarchand^35^, Núria Malats^36^, Roger Milne^37,38,39^, Steven C Moore^2^, Rachel E. Neale^40^, Kimmie Ng^41^, Ann L. Oberg^6^, Irene Orlow^42^, Alpa V Patel^43^, Ulrike Peters^44^, Miquel Porta^45^, Kari G. Rabe^46^, Francisco X Real^47,48^, Harvey A. Risch^49^, Nathaniel Rothman^10^, Paul Scheet^50^, Howard D Sesso^18,14^, Veronica W Setiawan^51^, Xiao-Ou Shu^52^, Debra Silverman^10^, Mingyang Song^14,53,54^, Meir J Stampfer^14,15,16^, Melissa C Southey^37,38,39^, Connie Thurman^14^, Geoffrey S Tobias^10^, Therese Truong^55^, Caroline Um^43^, Kala Visvanathan^56,57^, Nicolas Wentzensen^10^, Jean Wactawski-Wende^58^, Herbert Yu^59^, Chen Yuan^41^, Wei Zheng^52^, Brian M Wolpin^41^, Rachael Z Stolzenberg-Solomon^10^, Alison P. Klein^60,61^, Laufey T Amundadottir^1^

^1^Laboratory of Translational Genomics, Division of Cancer Epidemiology and Genetics, National Cancer Institute, National Institutes of Health, Bethesda, MD, USA, ^2^Metabolic Epidemiology Branch, Division of Cancer Epidemiology and Genetics, National Cancer Institute, National Institutes of Health, Bethesda, MD, USA, ^3^Occupational and Environmental Epidemiology Branch, Division of Cancer Epidemiology and Genetics, National Cancer Institute, National Institutes of Health, Bethesda, MD, USA, ^4^Department of Quantitative Health Sciences, Mayo Clinic College of Medicine, Jacksonville, FL, USA, ^5^Departments of Obstetrics and Gynecology and Population Health, NYU Grossman School of Medicine, NYU Perlmutter Comprehensive Cancer Center, New York, NY, USA, ^6^Department of Quantitative Health Sciences, Mayo Clinic College of Medicine, Rochester, MN, USA, ^7^Department of Epidemiology and Biostatistics, University of California San Francisco, San Francisco, CA, USA, ^8^Unit of Genetics., Department of Biology, University of Pisa, Pisa, Italy, ^9^Genomic Epidemiology Group, German Cancer Research Center (DKFZ), Heidelberg, Germany, ^10^Division of Cancer Epidemiology and Genetics, National Cancer Institute, National Institutes of Health, Bethesda, MD, USA, ^11^Department of Population Health, NYU Grossman School of Medicine, NYU Perlmutter Comprehensive Cancer Center, New York, NY, USA, ^12^Cancer Genomics Research Laboratory, Frederick National Lab for Cancer Research, Frederick, MD, USA, ^13^Department of Epidemiology and Biostatistics, Memorial Sloan Kettering Cancer Center,, New York, NY, USA, ^14^Department of Epidemiology, Harvard T.H. Chan School of Public Health, Boston, MA, USA, ^15^Channing Division of Network Medicine, Department of Medicine, Brigham and Women’s Hospital and Harvard Medical School, Boston, MA, USA, ^16^Department of Nutrition, Harvard T. H. Chan School of Public Health, Boston, MA, USA, ^17^Lunenfeld-Tanenbaum Research Institute, Sinai Health System and University of Toronto,, Toronto, Ontario, Canada, ^18^Division of Preventive Medicine, Brigham and Women’s Hospital, Boston, MA, USA, ^19^Division of Aging, Brigham and Women’s Hospital, Boston, MA, USA, ^20^Department of Epidemiology, Columbia University, New York, NY, USA, ^21^Department of Pathology, Johns Hopkins School of Medicine, Baltimore, MD, USA, ^22^SWOG Statistical Center, Fred Hutchinson Cancer Center, Seattle, WA, USA, ^23^Department of Preventive Medicine, Keck School of Medicine, University of Southern California, Los Angeles, CA, USA, ^24^Department of Epidemiology, Cancer Prevention, The University of Texas MD Anderson Cancer Center, Houston, TX, USA, ^25^Prosserman Centre for Population Health Research, Lunenfeld-Tanenbaum Research Institute, Sinai Health System, Toronto, Ontario, Canada, ^26^Division of Cancer Epidemiology, German Cancer Research Center (DKFZ), Heidelberg, Germany, ^27^ISGlobal, Centre for Research in Environmental Epidemiology (CREAL), Barcelona, Spain, ^28^Hospital del Mar Institute of Medical Research (IMIM), Universitat Autònoma de Barcelona, Barcelona, Spain, ^29^Division of Public Health Sciences, Fred Hutchinson Cancer Center, Seattle, WA, USA, ^30^Division of Environmental Epidemiology and Veterinary Public Health, Institute for Risk Assessment Sciences, Utrecht University, Utrecht, The Netherlands, ^31^Julius Global Health, The Julius Center for Health Sciences and Primary Care, University Medical Center, ^32^Trans-Divisional Research Program (TDRP), Division of Cancer Epidemiology and Genetics, National Cancer Institute, National Institutes of Health, Bethesda, MD, USA, ^33^Gastroenterology, Hepatology, and Nutrition Service, Memorial Sloan Kettering Cancer Center, New York, NY, USA, ^34^Division of Preventive Medicine, Department of Medicine, Brigham and Women’s Hospital, Boston, MA, USA, ^35^Population Sciences in the Pacific Program, Cancer Epidemiology, University of Hawaii Cancer Center, Honolulu, HI, USA, ^36^Genetic and Molecular Epidemiology Group, Spanish National Cancer Research Center (CNIO), Madrid, Spain, ^37^Cancer Epidemiology Division, Cancer Council Victoria, Melbourne, VIC, Australia, ^38^Centre for Epidemiology and Biostatistics, Melbourne School of Population and Global Health, The University of Melbourne, Melbourne, VIC, Australia, ^39^Precision Medicine, School of Clinical Sciences at Monash Health, Monash University, Clayton, VIC, Australia, ^40^Department of Population Health, QIMR Berghofer Medical Research Institute, Queensland, Australia, ^41^Department of Medical Oncology, Dana-Farber Cancer Institute, Harvard Medical School, Harvard University, Boston, MA, USA, ^42^Department of Epidemiology and Biostatistics, Memorial Sloan Kettering Cancer Center, New York, NY, USA, ^43^Department of Population Science, American Cancer Society, Atlanta, GA, USA, ^44^Division of Public Health Sciences, Fred Hutchinson Cancer Center, Seattle, WA, USA, ^45^Hospital del Mar Institute of Medical Research (IMIM), Centro de Investigacion Biomedica en Red de Epidemiologia y Salud Publica (CIBERESP), Barcelona, Spain, ^46^Department of Quantitative Health Sciences Research, Mayo Clinic College of Medicine, Rochester, MN, USA, ^47^Epithelial Carcinogenesis Group, Tumor Biology Programme, Spanish National Cancer Research Center (CNIO), Madrid, Spain, ^48^Department of Medicine and Life Sciences, Universitat Pompeu Fabra, Barcelona, Spain, ^49^Department of Chronic Disease Epidemiology, Yale School of Public Health, New Haven, CT, USA, ^50^Dept of Epidemiology, The University of Texas MD Anderson Cancer Center, Houston, TX, USA, ^51^Department of Population and Public Health Sciences, Keck School of Medicine, University of Southern California, Los Angeles, CA, USA, ^52^Division of Epidemiology, Department of Medicine, Vanderbilt Epidemiology Center, Vanderbilt-Ingram Cancer Center, Vanderbilt University School of Medicine, Nashville, TN, USA, ^53^Department of Nutrition, Harvard T.H. Chan School of Public Health, Boston, MA, USA, ^54^Clinical and Translational Epidemiology Unit and Division of Gastroenterology, Massachusetts General Hospital and Harvard Medical School, Boston, MA, USA, ^55^Paris-Saclay University, UVSQ, Inserm, Gustave Roussy, CESP, Villejuif, France, ^56^Department of Epidemiology, Johns Hopkins School of Public Health, Baltimore, MD, USA, ^57^Department of Oncology, Sidney Kimmel Comprehensive Cancer Center, Johns Hopkins School of Medicine, Baltimore, MD, USA, ^58^Department of Epidemiology and Environmental Health, University of Buffalo, Buffalo, NY, USA, ^59^Epidemiology Program, University of Hawaii Cancer Center, Honolulu, HI, USA, ^60^Department of Oncology, Sidney Kimmel Comprehensive Cancer Center, Johns Hopkins School of Medicine, Baltimore, MD, USA, ^61^Department of Medicine, Division of Gastroenterology & Hepatology, Johns Hopkins School of Medicine, Baltimore, MD, USA.

## References

1. Alsheikh, A. J. et al. The landscape of GWAS validation; systematic review identifying 309 validated non-coding variants across 130 human diseases. BMC Med Genomics 15, 74 (2022).

2. Lappalainen, T. et al. Transcriptome and genome sequencing uncovers functional variation in humans. Nature 501, 506–511 (2013).

3. Aguet, F. et al. Genetic effects on gene expression across human tissues. Nature 550, 204–213 (2017).

4. Wang, Z. et al. Imputation and subset-based association analysis across different cancer types identifies multiple independent risk loci in the TERT-CLPTM1L region on chromosome 5p15.33. Human Molecular Genetics 23, 6616–6633 (2014).

5. Chen, H. et al. Large-scale cross-cancer fine-mapping of the 5p15.33 region reveals multiple independent signals. Human Genetics and Genomics Advances 2, 100041 (2021).

6. Bhattacharjee, S. et al. A Subset-Based Approach Improves Power and Interpretation for the Combined Analysis of Genetic Association Studies of Heterogeneous Traits. The American Journal of Human Genetics 90, 821–835 (2012).

7. Fang, J. et al. Functional characterization of a multi-cancer risk locus on chr5p15.33 reveals regulation of TERT by ZNF148. Nature Communications 8, 1–15 (2017).

8. Kim, N. W. et al. Specific Association of Human Telomerase Activity with Immortal Cells and Cancer. Science 266, 2011–2015 (1994).

9. Castelo-Branco, P. et al. Methylation of the TERT promoter and risk stratification of childhood brain tumours: an integrative genomic and molecular study. The Lancet Oncology 14, 534–542 (2013).

10. Shay, J. W. & Bacchetti, S. A survey of telomerase activity in human cancer. European Journal of Cancer 33, 787–791 (1997).

11. Yoo, S. S., et al. *TERT* Polymorphism rs2853669 Influences on Lung Cancer Risk in the Korean Population. J Korean Med Sci 30, 1423 (2015).

12. Bojesen, S. E. et al. Multiple independent variants at the TERT locus are associated with telomere length and risks of breast and ovarian cancer. Nat Genet 45, 371–384 (2013).

13. Beesley, J. et al. Functional Polymorphisms in the TERT Promoter Are Associated with Risk of Serous Epithelial Ovarian and Breast Cancers. PLOS ONE 6, e24987 (2011).

14. Walsh, K. M. et al. Variants near TERT and TERC influencing telomere length are associated with high-grade glioma risk. Nat Genet 46, 731–735 (2014).

15. Mosrati, M. A. et al. TERT promoter mutations and polymorphisms as prognostic factors in primary glioblastoma. Oncotarget 6, 16663–16673 (2015).

16. Harland, M. et al. Germline TERT promoter mutations are rare in familial melanoma. Familial Cancer 15, 139–144 (2016).

17. George, J. R. et al. Association of TERT Promoter Mutation, But Not BRAF Mutation, With Increased Mortality in PTC. J Clin Endocrinol Metab 100, E1550–E1559 (2015).

18. Rachakonda, P. S. et al. TERT promoter mutations in bladder cancer affect patient survival and disease recurrence through modification by a common polymorphism. Proceedings of the National Academy of Sciences 110, 17426–17431 (2013).

19. Pópulo, H. et al. TERT promoter mutations in skin cancer: the effects of sun exposure and X-irradiation. J Invest Dermatol 134, 2251–2257 (2014).

20. Griewank, K. G. et al. TERT Promoter Mutation Status as an Independent Prognostic Factor in Cutaneous Melanoma. J Natl Cancer Inst 106, dju246 (2014).

21. Spiegl-Kreinecker, S. et al. Prognostic quality of activating TERT promoter mutations in glioblastoma: interaction with the rs2853669 polymorphism and patient age at diagnosis. Neuro Oncol 17, 1231–1240 (2015).

22. Horn, S. et al. TERT promoter mutations in familial and sporadic melanoma. Science 339, 959–961 (2013).

23. Hosen, I. et al. Mutations in TERT promoter and FGFR3 and telomere length in bladder cancer. International Journal of Cancer 137, 1621–1629 (2015).

24. Labussière, M. et al. TERT promoter mutations in gliomas, genetic associations and clinico-pathological correlations. Br J Cancer 111, 2024–2032 (2014).

25. Hiyama, E. et al. Telomerase activity is detected in pancreatic cancer but not in benign tumors. Cancer Res 57, 326–331 (1997).

26. Bell, R. J. A. et al. Understanding TERT Promoter Mutations: a Common Path to Immortality. Mol Cancer Res 14, 315–323 (2016).

27. James, M. A., Vikis, H. G., Tate, E., Rymaszewski, A. L. & You, M. CRR9/CLPTM1L Regulates Cell Survival Signaling and Is Required for Ras Transformation and Lung Tumorigenesis. Cancer Res 74, 1116–1127 (2014).

28. Clarke, W. R., Amundadottir, L. & James, M. A. CLPTM1L/CRR9 ectodomain interaction with GRP78 at the cell surface signals for survival and chemoresistance upon ER stress in pancreatic adenocarcinoma cells. International Journal of Cancer 144, 1367–1378 (2019).

29. Jia, J. et al. CLPTM1L Promotes Growth and Enhances Aneuploidy in Pancreatic Cancer Cells. Cancer Res 74, 2785–2795 (2014).

30. Parashar, D. et al. Targeted biologic inhibition of both tumor cell-intrinsic and intercellular CLPTM1L/CRR9-mediated chemotherapeutic drug resistance. npj Precis. Onc. 5, 16 (2021).

31. James, M. A. et al. Functional Characterization of CLPTM1L as a Lung Cancer Risk Candidate Gene in the 5p15.33 Locus. PLOS ONE 7, e36116 (2012).

32. Derrien, A.-C. et al. Functional characterization of 5p15.33 risk locus in uveal melanoma reveals rs452384 as a functional variant and NKX2.4 as an allele-specific interactor. Am J Hum Genet 109, 2196–2209 (2022).

33. Chang, Y.-H. et al. Isoform-level analyses of 6 cancers uncover extensive genetic risk mechanisms undetected at the gene-level. Br J Cancer 133, 874–885 (2025).

34. Zhong, J. et al. A Transcriptome-Wide Association Study Identifies Novel Candidate Susceptibility Genes for Pancreatic Cancer. JNCI: Journal of the National Cancer Institute 10.1093/jnci/djz246 (2020) doi:10.1093/jnci/djz246.

35. Yang, Y.-C. et al. rs401681 and rs402710 confer lung cancer susceptibility by regulating TERT expression instead of CLPTM1L in East Asian populations. Carcinogenesis 39, 1216–1221 (2018).

36. Wei, R. et al. TERT Polymorphism rs2736100-C Is Associated with EGFR Mutation-Positive Non-Small Cell Lung Cancer. Clin Cancer Res 21, 5173–5180 (2015).

37. Helbig, S. et al. Functional dissection of breast cancer risk-associated TERT promoter variants. Oncotarget 8, 67203–67217 (2017).

38. Ge, M. et al. Functional evaluation of TERT-CLPTM1L genetic variants associated with susceptibility of papillary thyroid carcinoma. Sci Rep 6, 26037 (2016).

39. Zhou, L. et al. The identification of two regulatory ESCC susceptibility genetic variants in the TERT-CLPTM1L loci. Oncotarget 7, 5495–5506 (2015).

40. Hsiung, C. A. et al. The 5p15.33 Locus Is Associated with Risk of Lung Adenocarcinoma in Never-Smoking Females in Asia. PLoS Genet 6, e1001051 (2010).

41. Florez-Vargas, O. et al. Genetic regulation of TERT splicing affects cancer risk by altering cellular longevity and replicative potential. Nat Commun 16, 1676 (2025).

42. Hofer, P. et al. MNS16A tandem repeats minisatellite of human telomerase gene: a risk factor for colorectal cancer. Carcinogenesis 32, 866–871 (2011).

43. Wang, L. et al. Association of a functional tandem repeats in the downstream of human telomerase gene and lung cancer. Oncogene 22, 7123–7129 (2003).

44. Yuan, H. et al. MNS16A polymorphism of the TERT gene on cancer risk: a systematic review and meta-analysis. Nucleosides, Nucleotides & Nucleic Acids 42, 1004–1018 (2023).

45. Zou, Y., Carbonetto, P., Wang, G. & Stephens, M. Fine-mapping from summary data with the “Sum of Single Effects” model. PLOS Genetics 18, e1010299 (2022).

46. Klein, A. P. et al. Genome-wide meta-analysis identifies five new susceptibility loci for pancreatic cancer. Nat Commun 9, 556 (2018).

47. Landi, M. T. et al. Genome-wide association meta-analyses combining multiple risk phenotypes provide insights into the genetic architecture of cutaneous melanoma susceptibility. Nat Genet 52, 494–504 (2020).

48. Koutros, S. et al. Genome-wide Association Study of Bladder Cancer Reveals New Biological and Translational Insights. Eur Urol 84, 127–137 (2023).

49. McKay, J. D. et al. Large-scale association analysis identifies new lung cancer susceptibility loci and heterogeneity in genetic susceptibility across histological subtypes. Nat Genet 49, 1126–1132 (2017).

50. Shi, J. et al. Genome-wide association study of lung adenocarcinoma in East Asia and comparison with a European population. Nat Commun 14, 3043 (2023).

51. Byrska-Bishop, M. et al. High-coverage whole-genome sequencing of the expanded 1000 Genomes Project cohort including 602 trios. Cell 185, 3426–3440.e19 (2022).

52. Wolpin, B. M. et al. Genome-wide association study identifies multiple susceptibility loci for pancreatic cancer. Nat Genet 46, 994–1000 (2014).

53. Zhong, J. et al. Large-scale multiomic analysis identifies non-coding somatic driver mutations and nominates ZFP36L2 as a driver gene for pancreatic ductal adenocarcinoma. Gut 10.1136/gutjnl-2025-335152 (2025) doi:10.1136/gutjnl-2025-335152.

54. Long, E. et al. High-throughput characterization of functional variants highlights heterogeneity and polygenicity underlying lung cancer susceptibility. Am J Hum Genet 111, 1405–1419 (2024).

55. Choi, J. et al. Massively parallel reporter assays of melanoma risk variants identify MX2 as a gene promoting melanoma. Nat Commun 11, 2718 (2020).

56. Colli, L. M. et al. Altered regulation of DPF3, a member of the SWI/SNF complexes, underlies the 14q24 renal cancer susceptibility locus. The American Journal of Human Genetics 108, 1590–1610 (2021).

57. Boyle, A. P. et al. Annotation of functional variation in personal genomes using RegulomeDB. Genome Res. 22, 1790–1797 (2012).

58. Breeze, C. E. et al. FORGEdb: a tool for identifying candidate functional variants and uncovering target genes and mechanisms for complex diseases. Genome Biol 25, 3 (2024).

59. Coetzee, S. G., Coetzee, G. A. & Hazelett, D. J. motifbreakR: an R/Bioconductor package for predicting variant effects at transcription factor binding sites. Bioinformatics 31, 3847–3849 (2015).

60. Tsherniak, A. et al. Defining a Cancer Dependency Map. Cell 170, 564–576.e16 (2017).

61. Li, W. et al. MAGeCK enables robust identification of essential genes from genome-scale CRISPR/Cas9 knockout screens. Genome Biol 15, 554 (2014).

62. Hsu, J. Y. et al. CRISPR-SURF: discovering regulatory elements by deconvolution of CRISPR tiling screen data. Nat Methods 15, 992–993 (2018).

63. Mukamel, R. E. et al. Repeat polymorphisms underlie top genetic risk loci for glaucoma and colorectal cancer. Cell 186, 3659–3673.e23 (2023).

64. Lee, O. W. et al. Targeted long-read sequencing of the Ewing sarcoma 6p25.1 susceptibility locus identifies germline-somatic interactions with EWSR1-FLI1 binding. The American Journal of Human Genetics 110, 427–441 (2023).

65. Grünewald, T. G. P. et al. Chimeric EWSR1-FLI1 regulates the Ewing sarcoma susceptibility gene EGR2 via a GGAA microsatellite. Nat Genet 47, 1073–1078 (2015).

66. Amundadottir, L. et al. Genome-wide association study identifies variants in the ABO locus associated with susceptibility to pancreatic cancer. Nat Genet 41, 986–990 (2009).

67. Petersen, G. M. et al. A genome-wide association study identifies pancreatic cancer susceptibility loci on chromosomes 13q22.1, 1q32.1 and 5p15.33. Nat Genet 42, 224–228 (2010).

68. Zagouri, F. et al. HTERT MNS16A polymorphism in breast cancer: a case–control study. Mol Biol Rep 39, 10859–10863 (2012).

69. Thomas-Chollier, M. et al. Transcription factor binding predictions using TRAP for the analysis of ChIP-seq data and regulatory SNPs. Nat Protoc 6, 1860–1869 (2011).

70. Yu, F.-X., Zhao, B. & Guan, K.-L. Hippo Pathway in Organ Size Control, Tissue Homeostasis, and Cancer. Cell 163, 811–828 (2015).

71. Landin Malt, A., et al. Alteration of TEAD1 Expression Levels Confers Apoptotic Resistance through the Transcriptional Up-Regulation of Livin. PLoS ONE 7, e45498 (2012).

72. Zhao, B., Li, L., Lei, Q. & Guan, K.-L. The Hippo–YAP pathway in organ size control and tumorigenesis: an updated version. Genes Dev. 24, 862–874 (2010).

73. Landin Malt, A., Georges, A., Silber, J., Zider, A. & Flagiello, D. Interaction with the Yes-associated protein (YAP) allows TEAD1 to positively regulate NAIP expression. FEBS Letters 587, 3216–3223 (2013).

74. Liu-Chittenden, Y. et al. Genetic and pharmacological disruption of the TEAD–YAP complex suppresses the oncogenic activity of YAP. Genes Dev. 26, 1300–1305 (2012).

75. Szeto, S. G. et al. YAP/TAZ Are Mechanoregulators of TGF-β-Smad Signaling and Renal Fibrogenesis. J Am Soc Nephrol 27, 3117–3128 (2016).

76. Weischer, M. et al. Short Telomere Length, Cancer Survival, and Cancer Risk in 47102 Individuals. J Natl Cancer Inst 105, 459–468 (2013).

77. Pooley, K. A. et al. A genome-wide association scan (GWAS) for mean telomere length within the COGS project: identified loci show little association with hormone-related cancer risk. Hum Mol Genet 22, 5056–5064 (2013).

78. Wentzensen, I. M., Mirabello, L., Pfeiffer, R. M. & Savage, S. A. The association of telomere length and cancer: a meta-analysis. Cancer Epidemiol Biomarkers Prev 20, 1238–1250 (2011).

79. Campa, D. et al. Leukocyte Telomere Length in Relation to Pancreatic Cancer Risk: A Prospective Study. Cancer Epidemiol Biomarkers Prev 23, 2447–2454 (2014).

80. Lynch, S. M. et al. A prospective analysis of telomere length and pancreatic cancer in the alpha-tocopherol beta-carotene cancer (ATBC) prevention study. International Journal of Cancer 133, 2672–2680 (2013).

81. Nan, H. et al. Shorter Telomeres Associate with a Reduced Risk of Melanoma Development. Cancer Res 71, 6758–6763 (2011).

82. Iles, M. M. et al. The Effect on Melanoma Risk of Genes Previously Associated With Telomere Length. J Natl Cancer Inst 106, dju267 (2014).

83. Seow, W. J. et al. Telomere Length in White Blood Cell DNA and Lung Cancer: A Pooled Analysis of Three Prospective Cohorts. Cancer Res 74, 4090–4098 (2014).

84. Antwi, S. O. et al. Leukocyte Telomere Length and Its Interaction with Germline Variation in Telomere-Related Genes in Relation to Pancreatic Adenocarcinoma Risk. Cancer Epidemiol Biomarkers Prev 29, 1492–1500 (2020).

85. Pearson, J. D. et al. Binary pan-cancer classes with distinct vulnerabilities defined by pro- or anti-cancer YAP/TEAD activity. Cancer Cell 39, 1115–1134.e12 (2021).

86. Jakubosky, D. et al. Properties of structural variants and short tandem repeats associated with gene expression and complex traits. Nat Commun 11, 2927 (2020).

87. Bhati, M., Mapel, X. M., Lloret-Villas, A. & Pausch, H. Structural variants and short tandem repeats impact gene expression and splicing in bovine testis tissue. Genetics 225, iyad161 (2023).

88. Jermusyk, A. et al. A 584 bp deletion in CTRB2 inhibits chymotrypsin B2 activity and secretion and confers risk of pancreatic cancer. Am J Hum Genet 108, 1852–1865 (2021).

89. Bakhtiari, M. et al. Variable number tandem repeats mediate the expression of proximal genes. Nat Commun 12, 2075 (2021).

90. Garg, P. et al. Pervasive *cis* effects of variation in copy number of large tandem repeats on local DNA methylation and gene expression. The American Journal of Human Genetics 108, 809–824 (2021).

91. Gymrek, M. et al. Abundant contribution of short tandem repeats to gene expression variation in humans. Nat Genet 48, 22–29 (2016).

92. Vinagre, J. et al. Frequency of TERT promoter mutations in human cancers. Nat Commun 4, 2185 (2013).

93. Coetzee, S. G. & Hazelett, D. J. motifbreakR v2: expanded variant analysis including indels and integrated evidence from transcription factor binding databases. Bioinformatics Advances 4, vbae162 (2024).

94. Murphy, A. E., Schilder, B. M. & Skene, N. G. MungeSumstats: a Bioconductor package for the standardization and quality control of many GWAS summary statistics. Bioinformatics 37, 4593–4596 (2021).

95. Chang, C. C. et al. Second-generation PLINK: rising to the challenge of larger and richer datasets. Gigascience 4, s13742-015-0047–8 (2015).

96. Machiela, M. J. & Chanock, S. J. LDlink: a web-based application for exploring population-specific haplotype structure and linking correlated alleles of possible functional variants: Fig. 1. Bioinformatics 31, 3555–3557 (2015).

97. Sanson, K. R. et al. Optimized libraries for CRISPR-Cas9 genetic screens with multiple modalities. Nat Commun 9, 5416 (2018).

98. PacificBiosciences/pbAA. PacBio (2025).

99. Dolzhenko, E. et al. Characterization and visualization of tandem repeats at genome scale. Nat Biotechnol 42, 1606–1614 (2024).

100. Poplin, R. et al. A universal SNP and small-indel variant caller using deep neural networks. Nat Biotechnol 36, 983–987 (2018).

101. Holt, J. M. et al. HiPhase: jointly phasing small, structural, and tandem repeat variants from HiFi sequencing. Bioinformatics 40, btae042 (2024).

102. Delaneau, O., Zagury, J.-F., Robinson, M. R., Marchini, J. L. & Dermitzakis, E. T. Accurate, scalable and integrative haplotype estimation. Nat Commun 10, 5436 (2019).

103. Danecek, P. et al. Twelve years of SAMtools and BCFtools. GigaScience 10, giab008 (2021).

104. Browning, B. L., Zhou, Y. & Browning, S. R. A One-Penny Imputed Genome from Next-Generation Reference Panels. The American Journal of Human Genetics 103, 338–348 (2018).

105. Zhang, Y. et al. Multisample motif discovery and visualization for tandem repeats. Genome Res. 35, 850–862 (2025).

106. Li, W. et al. Quality control, modeling, and visualization of CRISPR screens with MAGeCK-VISPR. Genome Biology 16, 281 (2015).

